# Serum methylation of *GALNT9*, *UPF3A*, *WARS,* and *LDB2* as non-invasive biomarkers for the early detection of colorectal cancer and premalignant adenomas

**DOI:** 10.1101/2021.11.11.21266182

**Authors:** María Gallardo-Gómez, Mar Rodríguez-Girondo, Núria Planell, Sebastian Moran, Luis Bujanda, Ane Etxart, Antoni Castells, Francesc Balaguer, Rodrigo Jover, Manel Esteller, Joaquín Cubiella, David Gómez-Cabrero, Loretta De Chiara

**Affiliations:** CINBIO, Universidade de Vigo, Vigo, Spain; Department of Biochemistry, Genetics and Immunology. Universidade de Vigo, Vigo, Spain; Galicia Sur Health Research Institute (IIS Galicia Sur), SERGAS-UVIGO, 36213 Vigo, Spain.; Department of Medical Statistics and Bioinformatics, Leiden University Medical Centre. Leiden, The Netherlands.; Translational Bioinformatics Unit, Navarrabiomed, Complejo Hospitalario de Navarra (CHN), Universidad Pública de Navarra (UPNA), IdiSNA, Pamplona, Spain; Cancer Epigenetics and Biology Program (PEBC), Bellvitge Biomedical Research Institute (IDIBELL). Avinguda de la Granvia, 199. 08908 L’Hospitalet de Llobregat, Barcelona, Spain; Department of Gastroenterology. Biodonostia Health Research Institute. Centro de Investigación Biomédica en Red de Enfermedades Hepáticas y Digestivas (CIBERehd). Universidad del País Vasco (UPV/EHU). San Sebastián, Spain; Department of Surgery. Hospital Universitario Donostia. San Sebastián. Spain; Gastroenterology Department, Hospital Clínic, IDIBAPS, CIBERehd, University of Barcelona. Barcelona, Spain; Department of Gastroenterology, Hospital General Universitario de Alicante. Alicante, Spain; Josep Carreras Leukaemia Research Institute (IJC), Badalona, Barcelona, Catalonia, Spain; Centro de Investigacion Biomedica en Red Cancer (CIBERONC), Madrid, Spain; Institucio Catalana de Recerca i Estudis Avançats (ICREA), Barcelona, Catalonia, Spain.; Physiological Sciences Department, School of Medicine and Health Sciences, University of Barcelona (UB), Barcelona, Catalonia, Spain.; Department of Gastroenterology, Complexo Hospitalario Universitario de Ourense, Instituto de Investigación Biomédica Galicia Sur, Centro de Investigación Biomédica en Red de Enfermedades Hepáticas y Digestivas (CIBERehd). Ourense, Spain; Biological & Environmental Sciences & Engineering Division, King Abdullah University of Science & Technology, Thuwal, Kingdom of Saudi Arabia.; Mucosal & Salivary Biology Division, King’s College London Dental Institute, London, SE1 9RT, United Kingdom.

**Keywords:** Advanced adenomas, Circulating cell-free DNA, Colorectal cancer, DNA methylation, Non-invasive biomarkers, Liquid biopsy, Screening, Serum.

## Abstract

**Background:** Early detection through screening programs has proven to be the most effective strategy to reduce the incidence and mortality of colorectal cancer. The most widely implemented non-invasive screening test is the fecal immunochemical test, which presents an inadequate sensitivity for the detection of precancerous advanced adenomas. This fact, together with the modest participation rates in screening programs, highlights the need for a blood test that could improve both the adherence to screening and the selection to colonoscopy.

**Methods:** In this study, we conducted a serum-based discovery and validation of circulating cell-free DNA (cfDNA) methylation biomarkers for colorectal cancer screening in a multicentre cohort of 433 serum samples including healthy controls, benign pathologies, advanced adenomas, and colorectal cancer. First, we performed an epigenome-wide methylation analysis with the MethylationEPIC array in 280 cfDNA samples using a pooling approach, followed by a robust prioritization of candidate biomarkers for the joint detection of advanced adenomas and colorectal cancer (advanced neoplasia). Then, candidate biomarkers were validated by pyrosequencing in independent individual 153 cfDNA samples.

**Results:** We report *GALNT9*, *UPF3A*, *WARS*, and *LDB2* as new non-invasive methylation biomarkers for the early detection of colorectal advanced neoplasia. A model composed of *GALNT9*, *UPF3A*, *WARS*, and *LDB2* reported a sensitivity of 62.1% and a specificity of 97.4% for the detection of advanced neoplasia. On the other hand, the combination of *GALNT9* and *UPF3A* by logistic regression discriminated advanced neoplasia with 78.8% sensitivity and 100% specificity, outperforming the commonly used fecal immunochemical test and the methylated *SEPT9* blood test.

**Conclusions:** Serum methylation levels of *GALNT9*, *UPF3A*, *WARS*, and *LDB2* represent highly specific and sensitive novel blood-based biomarkers for the detection of colorectal cancer and premalignant advanced adenomas of both distal and proximal locations. The reported results show the feasibility of DNA sample pooling strategies for biomarker discovery. Overall, this study highlights the utility of cfDNA methylation for the early detection of colorectal neoplasia, with the potential to be implemented as a non-invasive test for colorectal cancer screening.

## BACKGROUND

Colorectal cancer (CRC) is the third cancer with the highest incidence worldwide and the second leading cause of cancer death in both sexes [1]. Diagnosis at advanced symptomatic stages is responsible for low survival (14% for stage IV) compared to 90% five-year survival for stage I and II [2]. A recent decrease in CRC-related mortality and incidence has been reported, mainly due to the implementation of screening programs that enable early detection of preclinical CRC and the removal of precancerous colorectal adenomas [3–5]. Despite strong evidence supports the reduction of both CRC incidence and mortality related to screening [6, 7], the overall participation rate in stool-based screening programs using the fecal immunochemical test (FIT) followed by a confirmatory colonoscopy remains modest (49.5% in Europe and 43.8% worldwide) [4, 8]. Also, although FIT reports high specificity (90-94%) and convenient sensitivity (73-88%) for colorectal tumors, the sensitivity for the detection of AA is moderate to low (22-56%) [9–12]. Since the effectiveness of a screening test relies not only on the diagnostic performance of the test but also on its acceptance by the target population, test preference for CRC screening has been evaluated. A survey-based study reported as first choice a blood test over a stool one [13]; similarly, among screening-enrolled individuals who refused colonoscopy, 83% preferred a non-invasive blood-based test, 15% chose a fecal test and 3% refused any test [14]. Therefore, participation in screening programs could significantly improve by offering a non-invasive blood-based test.

Liquid biopsy has emerged as a non-invasive alternative to traditional procedures for sampling. Blood-based screening has the advantage of being easily available, repeatable, and minimally invasive for the patient [15]. Circulating cell-free DNA (cfDNA) can be detected in body fluids and has been proposed as a source of liquid biopsy biomarkers as it reflects alterations occurring during neoplastic transformation, such as aberrant methylation in colorectal carcinogenesis [16, 17]. Recent epigenome-wide methylation analyses have reported that alterations in DNA methylation arise during the early stages of tumor progression and that the heterogeneity of the different pathways to CRC is already detectable in colorectal adenomas [18, 19]. The Illumina MethylationEPIC BeadChip array has proven to be reliable and consistent for DNA methylation analysis [20], and its combination with sample pooling represents a particularly suitable strategy for the cost-effective analysis of large sample sets aiming to discover differentially methylated signatures [21, 22]. In this study, following a cfDNA pooling strategy, we aimed to identify non-invasive methylation biomarkers for the early detection of both colorectal cancer and advanced precancerous lesions. Here we report the discovery and independent validation of combined serum-based methylation biomarkers that provide a new highly specific and sensitive non-invasive test for the screening and early detection of colorectal cancer and advanced adenomas.

## METHODS

### Study design

The study was conducted in three phases: (i) a high-throughput discovery analysis, paired with a statistically robust biomarker prioritization, was performed to identify candidate non-invasive methylation biomarkers for CRC screening (joint detection of AA and CRC), using a sample pooling strategy. Next, targeted assays were designed and optimized for the quantification of the candidate biomarkers in an independent cohort of patients (individual serum samples). The targeted analysis was divided into (ii) a preliminary evaluation of the candidate biomarkers in a small subset of samples, followed by the application of penalized regression models to further reduce the number of biomarkers and to obtain specific predictive biomarkers subsets; and (iii) a subsequent validation and final statistical model construction in a larger serum sample set, based on the selected biomarkers. The final classification models were also evaluated in non-colorectal tumors. An overview of the study design is shown in Figure 1.

**Figure 1.**
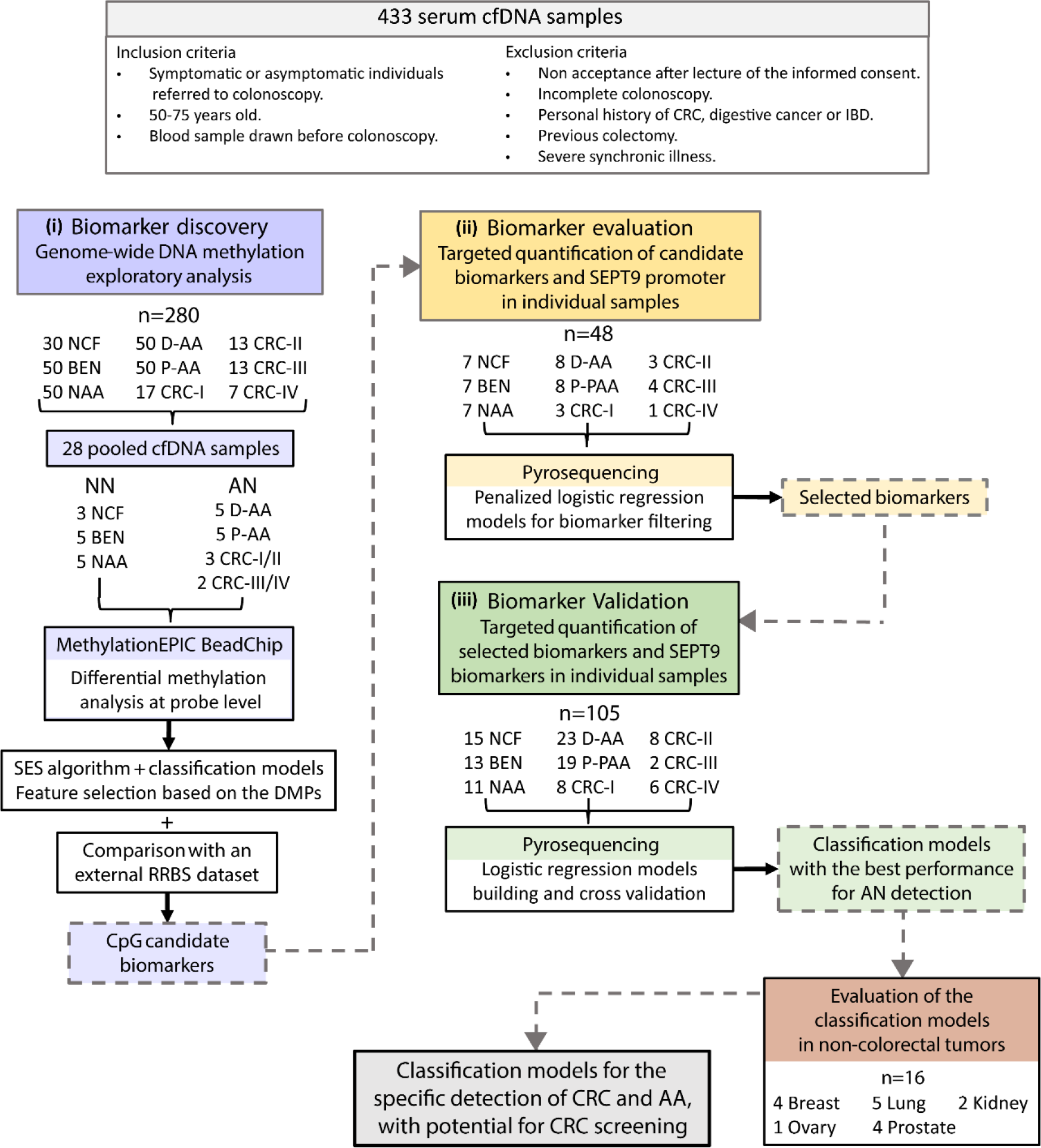
Study workflow. The study was divided into (i) a biomarker discovery phase, (ii) a candidate biomarker evaluation phase, and (iii) a selected biomarker validation. cfDNA: cell-free DNA; CRC: colorectal cancer (AJCC staging system); IBD: inflammatory bowel disease; NCF: no colorectal findings; BEN: benign pathology; NAA: non-advanced adenomas; D-AA: distal advanced adenomas; P-AA: proximal advanced adenomas; NN: no neoplasia; AN: advanced neoplasia; DMP: differentially methylated position; RRBS: reduced representation bisulfite sequencing.

### Patients and samples

Individuals were recruited from the following Spanish Hospitals: Complexo Hospitalario Universitario de Ourense (Ourense), Hospital Clínic de Barcelona (Barcelona), Hospital Donostia (San Sebastián), and Hospital General Universitario de Alicante (Alicante). A total of 435 individuals between 50-75 years old were selected. Exclusion criteria included a personal history of CRC, digestive cancer or inflammatory bowel disease, a severe synchronic illness, and a previous colectomy. All individuals included in the study underwent a colonoscopy which was performed by experienced endoscopists following the recommendations from the Spanish guidelines on quality of colonoscopy [23]. Blood samples were obtained immediately before the colonoscopy procedure. Blood samples were coagulated and subsequently centrifuged according to the manufacturer’s instructions for serum collection. Circulating cell-free DNA (cfDNA) was extracted from 0.5-2 mL serum according to availability. Serum samples were stored at −20 °C until cfDNA extraction.

Individuals were classified according to the most advanced colorectal finding. Lesions were considered ‘proximal’ when located only proximal to the splenic flexure of the colon and ‘distal’ when found only in the distal colon or in both distal and proximal colon. Advanced adenomas (AA) are defined as adenomas ≥ 1 cm, with villous components or high-grade dysplasia.

We performed a stratified random sampling using colorectal findings and sex as stratifying variables. Strata were matched by age and recruitment hospital. This multicenter cohort was separated into two independent subsets: Biomarker discovery sample set (n=280; 140 female and 140 male) and Biomarker validation sample set (n=153; 73 female and 80 male). A description of the independent cohorts is presented in Table 1.

**Table 1.** Independent cohorts used for discovery, evaluation, and validation of biomarkers. The number of patients, age median and range, sex, and colorectal findings are provided for each sub-cohort. AN: advanced neoplasia; NN: no neoplasia; NCF: no colorectal findings; BEN: benign pathology; NAA: non-advanced adenomas; D-AA: distal advanced adenomas; P-AA: proximal advanced adenomas; T: tubular; TV: tubulovillous; V: villous; LGD: low-grade dysplasia; HGD: high-grade dysplasia; CRC: colorectal cancer; AJCC: American Joint Committee on Cancer staging system.

|  |  | Discovery cohort<br>n=280 |  | Biomarker<br>evaluation cohort<br>n=48 |  | Biomarker<br>validation cohort<br>n=105 |  |
| --- | --- | --- | --- | --- | --- | --- | --- |
|  |  | AN | NN | AN | NN | AN | NN |
| Total (n) |  | 140 | 140 | 27 | 21 | 66 | 39 |
| Age median<br>(range) |  | 62.0<br>(51-72) | 62.0<br>(51-72) | 62.5<br>(50-75) | 62.0<br>(51-75) | 63.5<br>(50-75) | 63.0<br>(50-72) |
| **Sex** |  |  |  |  |  |  |  |
|  | Male | 70 | 70 | 14 | 10 | 36 | 20 |
|  | Female | 70 | 70 | 13 | 11 | 30 | 19 |
| **NCF** |  | - | **30** | - | **7** | - | **15** |
| **BEN** |  | - | **50** | - | **7** | - | **13** |
|  | Hemorrhoids | - | 25 | - | 3 | - | 8 |
|  | Diverticula | - | 25 | - | 4 | - | 5 |
| **NAA** |  | - | **50** | - | **7** | - | **11** |
| **AA** |  | **100** | - | **16** | - | **42** | - |
| AA histology |  |  | - |  |  |  |  |
|  | T (size >10mm) | 52 | - | 11 | - | 24 | - |
|  | TV | 36 | - | 3 | - | 15 | - |
|  | V | 12 | - | 2 | - | 3 | - |
| AA dysplasia |  |  |  |  |  |  |  |
|  | Without dysplasia | 7 | - | . | - | 5 | - |
|  | LGD | 89 | - | 13 | - | 31 | - |
|  | HGD | 4 | - | 3 | - | 7 | - |
| AA localization |  |  |  |  |  |  |  |
|  | Distal | 50 | - | 8 | - | 23 | - |
|  | Proximal | 50 | - | 8 | - | 19 | - |
| **CRC** |  | **50** | - | **11** | - | **24** | - |
| CRC AJCC stage |  |  |  |  |  |  |  |
|  | Stage I | 16 | - | 3 | - | 8 | - |
|  | Stage II | 14 | - | 3 | - | 8 | - |
|  | Stage III | 14 | - | 4 | - | 2 | - |
|  | Stage IV | 6 | - | 1 | - | 6 | - |
| CRC localization |  |  |  |  |  |  |  |
|  | Distal | 39 | - | 9 | - | 12 | - |
|  | Proximal | 11 | - | 2 | - | 12 | - |

Biomarker discovery sample set included 30 individuals with no colorectal findings (NCF), 50 with benign pathologies (BEN: hemorrhoids and diverticula), 50 with non-advanced adenomas (NAA), 50 with proximal AA (P-AA), 50 with distal AA (D-AA), and 50 CRC cases (17 stage I, 13 stage II, 13 stage III and 7 stage IV), according to the AJCC staging system [24]. On the other hand, the Biomarker evaluation and validation sample set comprised 22 NCF individuals, 20 BEN, 18 NAA, 31 D-AA, 27 P-AA, and 35 CRC cases (11 stage I, 11 stage II, 6 stage III, and 7 stage IV). ‘Advanced colorectal neoplasia’ (AN) was defined as AA or CRC. In contrast, NCF, BEN, and NAA were considered together as ‘no neoplasia’ (NN).

The specificity of the biomarkers for the detection of colorectal cancer and advanced adenomas was evaluated in an independent cohort of 16 patients with different cancer types, including breast (n=4), kidney (n=2), lung (n=5), ovary (n=1), and prostate (n=4) cancer (Supplementary Table 4). Additionally, 8 pairs of matched serum and plasma samples from the same individual were used to account for differences in the methylation levels between serum and plasma (3 NCF, 2 BEN, and 3 NAA).

### DNA extraction and sample pooling

cfDNA from samples used for biomarker discovery was extracted with a phenol-chloroform protocol as described by Hufnagl et al. [25], with minor modifications. DNA was quantified in each sample using Qubit dsDNA HS Assay Kit (Thermo Fisher Scientific, Waltham, MA, USA). For methylation biomarker discovery we followed a DNA pooling strategy. A detailed description of the pooling protocol can be found in [22]. 28 independent cfDNA pooled samples were constructed using equal amounts of cfDNA from 5 men and 5 women from the same pathological group, recruitment hospital- and age-matched. Pools were divided into no neoplasia (NN) and advanced colorectal neoplasia (AN). The NN group comprised 13 cfDNA pooled samples: 3 pools of NCF individuals, 5 pools of BEN (hemorrhoids and diverticula), and 5 pools of individuals with NAA. On the other hand, the 15 cfDNA pooled samples of advanced neoplasia (AN) included 5 pools of individuals with AA located only in the proximal colon (P-AA), 5 pools of individuals with AA located in the distal colon of both proximal and distal locations (D-AA), 3 pools of CRC patients stages I and II, and 2 pools of CRC stages III and IV (Supplementary Table 2). The cfDNA pools were stored at − 20 °C and were sent to the Cancer Epigenetics and Biology Program (PEBC) facilities at the Bellvitge Biomedical Research Institute (IDIBELL, Barcelona, Spain) for processing and methylation quantification.

The QIAmp Circulating Nucleic Acid Kit (Qiagen, Hilden, Germany) was used for cfDNA extraction from serum and plasma samples in the evaluation and validation phase. Individual cfDNA samples were bisulfite-converted using EZ DNA Methylation-Direct Kit (Zymo Research, Irvine, CA, USA) according to the manufacturer’s protocol. Bisulfite treated cfDNA samples were stored at -80°C.

### Genome-wide DNA methylation measurements

DNA methylation of pooled samples was measured with the Infinium MethylationEPIC BeadChip array (Illumina, San Diego, CA, USA). Pooled samples were bisulfite-treated and arrays were hybridized following the manufacturer’s instructions. A total of 865,859 CpG sites were quantified throughout the genome, covering promoter CpG islands, RefSeq genes, open chromatin, and enhancer intergenic regions identified by FANTOM5 and ENCODE projects [26–28]. Importantly, to minimize the potential impact of batch effects and confounder variability, samples of each pathological group were randomly allocated to the slides.

### Methylation biomarker discovery

Illumina methylation data were preprocessed and analyzed using the R environment (versions 3.3.3 and 3.4.0) [29] with R and Bioconductor [30] packages (see Additional File 2 information for details on quality control and preprocessing). Methylation levels were expressed as beta-values for visualization and intuitive interpretation of the results. Methylation expressed as M-values (*logit* transformation of the beta-values) were used for differential methylation analysis and biomarker selection, as recommended by Du et al. [31]. To test for differentially methylated positions (DMPs) between AN (AA or CRC) and NN (NCF, BEN, or NAA) we used the *limma* package [8]: linear models were fitted for each CpG site across all samples by generalized least squares, and an empirical Bayes method was used to compute the p-values. As recommended by Mansell et al., [32] linear regression assumptions were checked for each model using the *gvlma* package [33].

To select and prioritize the DMPs as candidate biomarkers we first applied the constraint-based Statistically Equivalent Signature (SES) algorithm for feature selection, contained in the *MXM* package [34]: multiple CpG sets with minimal size and maximal predictive power for the binary classification problem NN vs AN were obtained by iteratively comparing logistic regression models through a chi-square test. Then, the different CpG subsets were used to build classification models based on support vector machine (*e1071* package) [35], random forest (*randomForest* package) [36], and logistic regression. Models were cross-validated to select candidate CpG biomarkers with minimum prediction error for NN vs AN classification.

Methylation microarray data were compared with an external RRBS (Reduced Representation Bisulfite Sequencing) dataset targeting CpG-rich regions of 20 AA and 27 CRC cases, with matched adenoma/tumor, healthy mucosa, and serum samples of each patient; and 24 serum samples from healthy controls. A subset of CpG sites that reported concordance in methylation differences between NN and AN in both datasets were added to the biomarker list.

### Targeted methylation quantification by bisulfite pyrosequencing

The methylation levels of the candidate biomarkers, together with the v2 promoter region of the SEPT9 gene, were evaluated by bisulfite pyrosequencing in 153 individual serum samples. PCR and sequencing primers were designed with the PyroMark Assay Design software (version 2.0.1.15, Qiagen, Hilden, Germany). Bisulfite-converted cfDNA (2 μl) was subjected to PCR amplification using primers flanking the CpG candidate biomarkers. Multiplex reactions including 3-6 candidate markers were performed, followed by nested singleplex PCR reactions using a biotin-labeled primer. Primers and PCR conditions for multiplex and singleplex PCR are provided in the supplementary information (see Additional File 2 and Supplementary Table 3). Biotin-labeled amplicons were further resolved and visualized on 3% agarose gels. Pyrosequencing was performed using a PyroMark Q96 ID pyrosequencer (Qiagen, Hilden, Germany) according to the manufacturer’s protocol for CpG methylation quantification. Data acquisition and methylation measurements were conducted at the Biomedical Research Institute of Malaga facilities (IBIMA, Málaga, Spain) using PyroMark Q96 ID software, CpG analysis mode (version 1.0.11).

Fully methylated and unmethylated controls were included in each run and were used to check bisulfite conversion, the performance of the assay, and to account for batch-to-batch variation. DNA extracted from peripheral blood from a donor was used to prepare controls. For the fully methylated control DNA was treated with CpG methyltransferase (M.SssI; New England Biolabs, Ipswich, MA, USA), while for the unmethylated control whole genome amplification was performed to eliminate methylation marks (illustra GenomiPhi V2 DNA Amplification Kit, GE Healthcare; Chicago, IL, USA).

### Biomarker selection and classification model development

Both raw and log10-transformed methylation percentages were subjected to multivariate analyses for the development and validation of methylation-based classification models. The validation set of 153 individual samples was randomly divided for the multi-step process of methylation panel development (Table 1).

First, a preliminary evaluation of the methylation levels of the candidate biomarkers was carried out in a 30% sample subset (n=48, 21 NN, 8 D-AA, 8 P-AA, and 11 CRC cases). Penalized logistic regressions Least Absolute Shrinkage and Selection Operator (LASSO) and Elastic net were applied to the candidate biomarkers, age, and sex for feature selection, with the *glmnet* R package [37]. The minimum mean cross-validation error was used to define the penalty factor for LASSO and Elastic net models. Biomarkers present in the LASSO and Elastic net-derived models were selected for validation. Then, multivariate logistic regressions were fitted in the remaining 70% samples (n=105, 39 NN, 23 D-AA, 19 P-AA, and 24 CRC cases) to derive models based on the selected biomarkers.

### Statistical analyses

All statistical analyses were performed with the R environment (version 3.4.0). In the epigenome-wide methylation analyses, *p*-values were adjusted for multiple testing with the Benjamini-Hochberg procedure to control the false discovery rate (FDR). One-sided Fisher’s exact tests were used to assess the significance of the enrichment of the DMPs to functionally annotated elements using the annotation of the complete array as background. To assess the performance of the classification models, receiver-operating characteristic (ROC) curves were elaborated, derived by the leave-one-out cross-validation approach, and AUC, sensitivity, and specificity values were estimated with their corresponding 95% confidence intervals. The best cut-off values were determined by the Youden Index method [38]. Negative and positive predictive values (NPV, PPV) were also estimated for the best cut-off values. Fisher’s exact tests were employed to compare the proportion of distal and proximal lesions detected. Wilcoxon rank-sum test was used to compare the methylation levels between NN and AN in individual serum samples. Non-parametrical Wilcoxon signed-rank test was used to compare methylation levels between matched serum and plasma samples.

## RESULTS

### Sample pooling

A total of 28 cfDNA pooled samples were used in the epigenome-wide methylation analysis for biomarker discovery. As described in [22], 10 individuals with the same colorectal pathology were included in each pool and were matched by recruitment hospital, sex, and age (median 62, range 51-72). The final quantity of cfDNA in the pools ranged from 62 to 403 ng. There was no statistically significant difference in the mean age between pools (ANOVA, *p*-value < 0.05). The age range matches the USPSTF guideline recommendation for CRC screening (50-75 years) [39]. The NN group comprised 13 cfDNA pooled samples (3 NCF, 5 BEN, and 5 NAA), while the AN included 15 cfDNA pooled samples (5 D-AA, 5 P-AA, 3 CRC stages I/II, and 2 CRC stages III/IV). A detailed description of the pooled samples can be found in Supplementary Table 2.

### Epigenome-wide biomarker discovery

MethylationEPIC BeadChip was used for quantitative DNA methylation profiling in the 28 cfDNA pooled samples. We correctly detected 97.78% of the total probes present on the array. All failed positions (1,795 probes with a detection *p*-value > 0.01 and 17,452 probes with a bead count < 3) were removed before analysis. After quality control and data preprocessing, 18,035 probes were discarded as they did not hold the assumptions for linear regression model fitting (i.e. linearity, homoscedasticity, uncorrelatedness, and normality of the standardized residuals). After quality control and preprocessing, a total of 741,310 CpG sites mapped to the human genome assembly GRCh37/hg19 were left for differential methylation testing. No samples were removed due to quality issues.

The purpose of screening programs includes the early detection of preclinical CRC and the detection and removal of AA [3, 40]; therefore, differential methylation was assessed between NN and AN groups. The analysis between these cfDNA pools revealed 376 differentially methylated positions (10% FDR, BH-adjusted *p*-value) (Fig. 2A), annotated to a total of 290 gene regions and 183 CpG islands. Chromosomes 1 and 2 hold the majority of DMPs (10.4% and 7.7%, respectively) possibly due to their larger length. Most CpG sites (326

**Figure 2.**
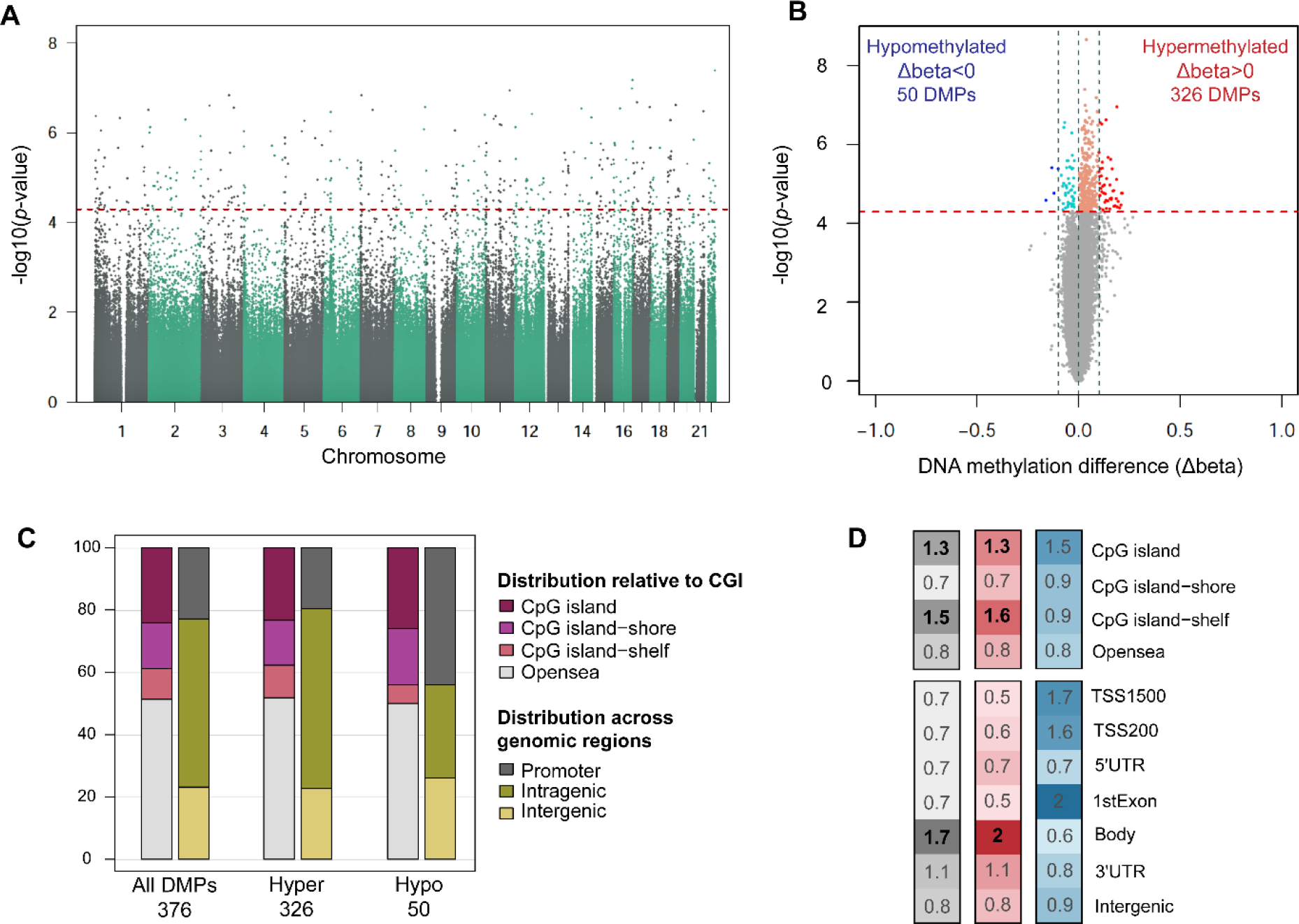
Distribution and annotation of differentially methylated positions (DMPs) between advanced neoplasia and no neoplasia pools. **A.** Manhattan plot for differential methylation. The -log10(*p*-value) for the 741,310 probes analyzed are sorted by chromosome location. Significant DMPs (376) appear above the red dashed line (FDR 10%) **B.** Volcano plot showing the -log10(*p*-value) versus differences in methylation levels (Δbeta: obtained by subtracting the DNA methylation levels (beta-values) of NN from AN). Significant hypermethylated (Δbeta > 0) and hypomethylated (Δbeta < 0) positions appear highlighted in color and above the red dashed line (FDR 10%). **C.** Distribution of the DMPs relative to CpG islands and functional genomic locations. **D.** Enrichment of DMPs in relation to CpG island annotation and functional genomic regions. The color scale indicates the fold enrichment of all DMPs (grey), hypermethylated (red), and hypomethylated (blue) positions. The bolded numbers indicate annotations that are enriched with respect to the distribution of probes on the MethylationEPIC array (one-sided Fisher’s exact test *p*-value < 0.05). Functional characterization of probes according to the Methylation EPIC Manifest (R package version 0.6.0) [41]: CpG island: region of at least 200bp with a CG content > 50% and an observed-to-expected CpG ratio ≥ 0.6; CpG island-shore: sequences 2 kb flanking the CpG island, CpG island-shelf: sequences 2 kb flanking shore regions, opensea: sequences located outside these regions, promoter regions (5′UTR, TSS200, TSS1500, and first exons), intragenic regions (gene body and 3′UTR), and intergenic regions. TSS200, TSS1500: 200 and 1500 bp upstream the transcription start site, respectively.

DMPs, 86.7%) were found to be hypermethylated in AN (Fig. 2B). Concerning the distribution across functional elements, DMPs were mainly located in open sea regions (51.59%) and CpG islands (23.67%) (Fig. 2C). Differentially hypermethylated positions were significantly enriched in CpG islands, shelves, and gene body regions (Fig. 2D). Clustering analyses of all pooled samples based on the methylation values of the 376 DMPs (Fig. 3A) suggest the capacity of this differentially methylated signature to discriminate advanced neoplasia from no neoplasia controls.

**Figure 3.**
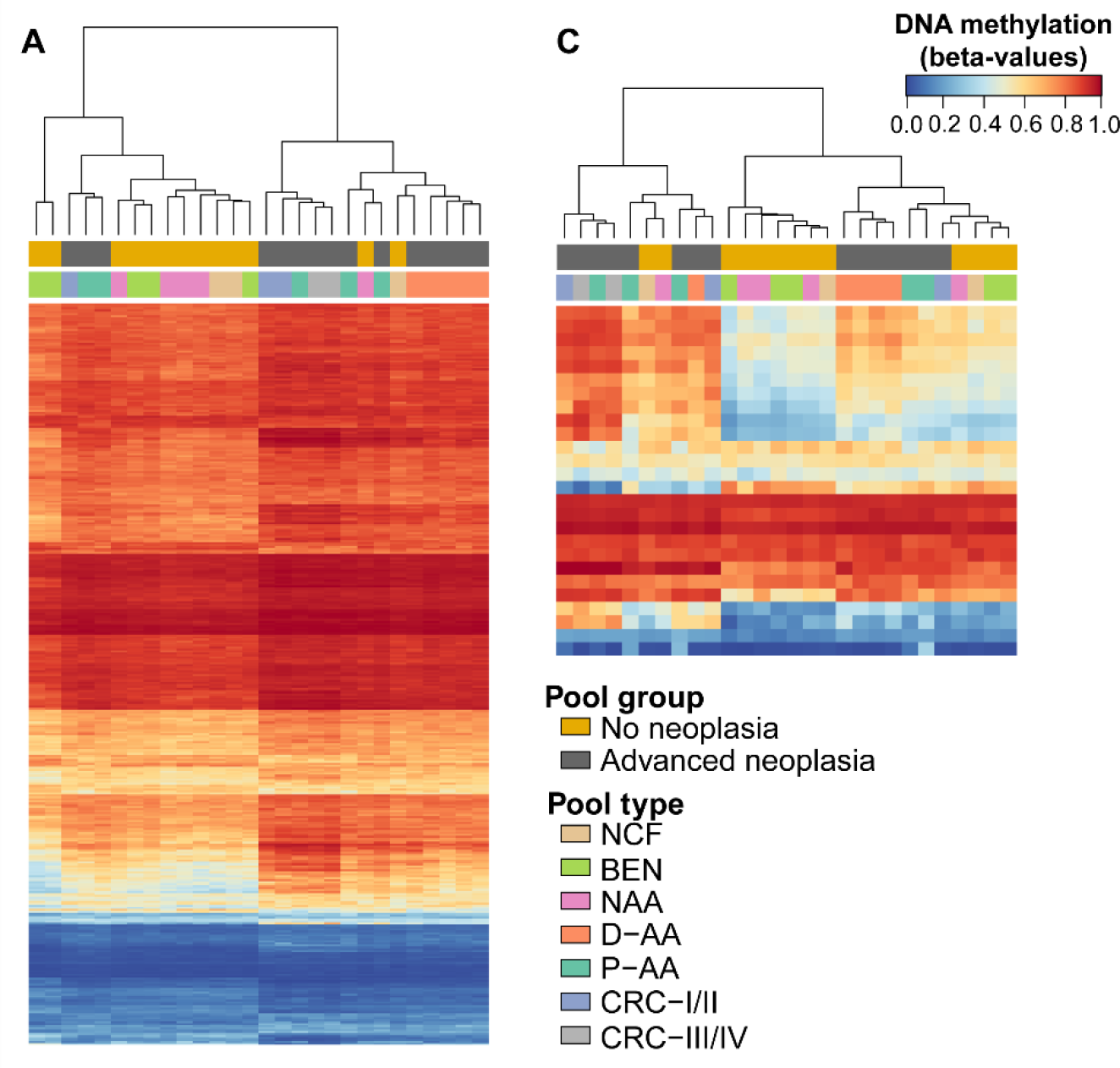
Unsupervised clustering analyses of the 28 cfDNA pooled samples. **A.** Hierarchical clustering and heatmap showing the methylation levels across all samples for the 376 DMPs. **B.** Hierarchical clustering and heatmap showing the methylation levels across all samples for the 26 candidate biomarkers. Each column represents one pool while rows correspond to CpG sites. Dendrograms were computed and reordered using Euclidean distance and a complete clustering agglomeration. Methylation levels are expressed as beta-values ranging from 0 (blue, unmethylated) to 1 (red, fully methylated).

In order to identify the most relevant features, we followed a robust strategy of selection. We first applied the constraint-based SES feature selection algorithm [34] on the 376 DMPs to identify combinations of CpGs whose performances for the NN vs AN classification were statistically equivalent. A total of 3,256 combinations of CpG pairs were obtained. Secondly, we used two strategies for filtering: (a) the CpG pairs were used to build classification models (support vector machine, logit regression, and random forest) that were cross-validated in the cfDNA pools by the leave-one-out strategy, due to limited sample size. CpG sites present in models with more than 20% prediction error were discarded. And then (b) the remaining CpG sites were ranked according to the mean difference in the methylation levels. From the ranked list we selected the top 15 CpG sites with greater methylation differences and present in models with minimum classification error. Finally, due to the limited sensitivity of FIT for the detection of advanced adenomas, especially those of proximal location, we also selected 3 additional CpG sites that presented 0% prediction error for the detection of advanced adenomas (1 CpG site derived from the NN vs D-AA comparison; 2 CpG sites derived from the NN vs P-AA comparison).

In parallel, we carried out a comparison between an external RRBS dataset and our methylation microarray data. The RRBS dataset comprised serum and tissue samples from patients with CRC, AA, and healthy controls. We combined both datasets to select a subset of 8 CpG sites that reported more than 30% differences in the methylation levels by bisulfite sequencing, and whose methylation differences showed the same direction in our methylation microarray data (i.e. hyper/hypomethylated in AN in both datasets).

Altogether, a total of 26 CpG positions were selected as candidate biomarkers whose methylation levels and their ability to discriminate colorectal advanced neoplasia are shown in Figure 3B. Three of 26 markers were hypomethylated, while the rest were found hypermethylated in AN compared to no neoplasia (Fig 4A). Description, regulatory features, and relation to CpG island of these 26 CpG candidate biomarkers are available in Supplementary Table 4.

**Figure 4.**
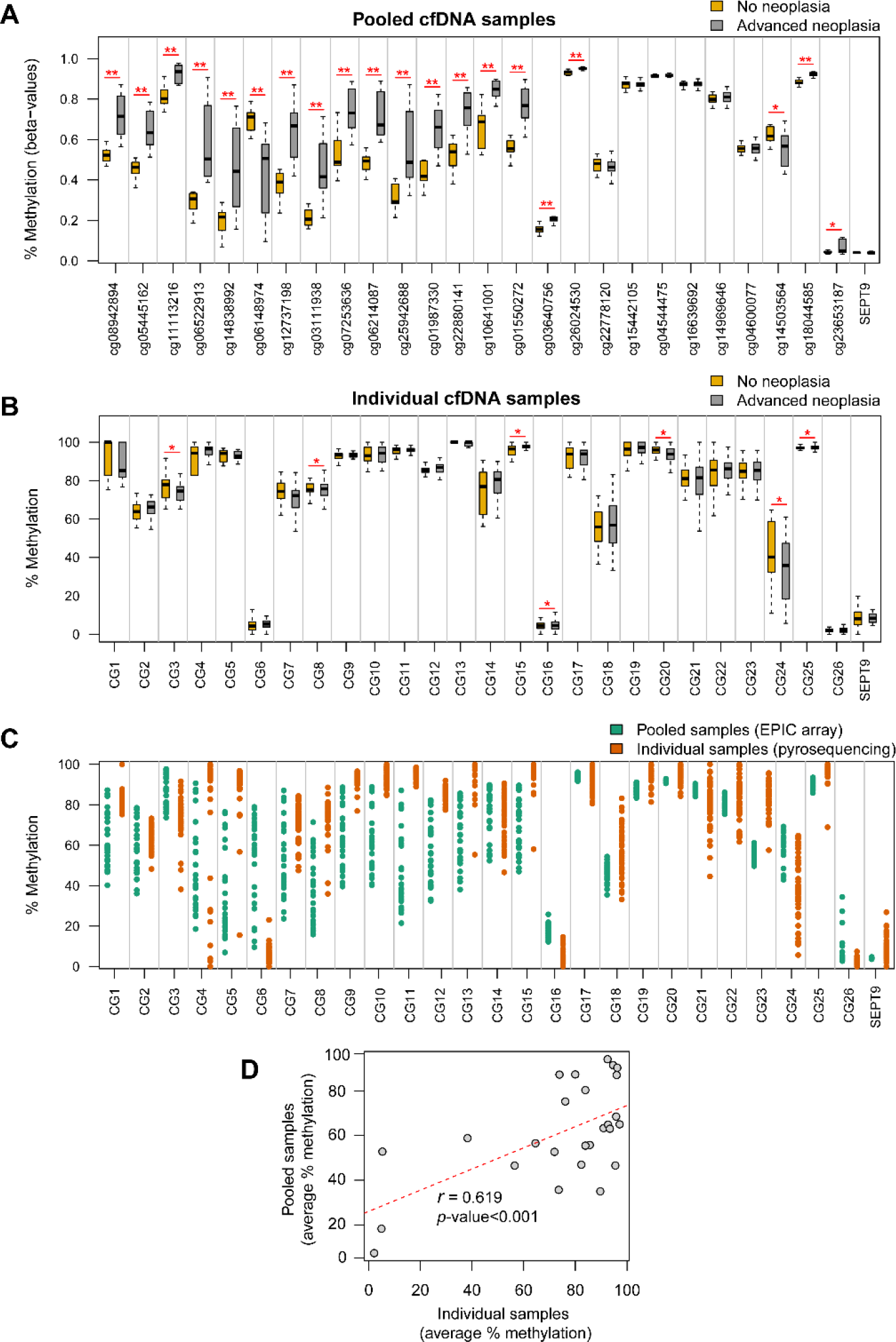
Methylation levels of the 26 candidate biomarkers. **A.** Methylation levels of the 26 CpG sites selected as candidate biomarkers in the 28 pooled cfDNA samples, with the corresponding MethylationEPIC CpG probe ID. Average methylation of the probes targeting the v2 promoter region of the SEPT9 gene (cg02884239, cg20275528, and cg12783819) is also shown. Methylation is shown as beta-values ranging from 0-unmethylated to 1-fully methylated (**differential methylation *p*-value < 0.01; *differential methylation *p*-value < 0.05). **B.** Methylation levels of the 26 candidate CpG sites and SEPT9 promoter in the biomarker evaluation cohort (n=48) of individual serum cfDNA samples. Methylation percentage was obtained through bisulfite pyrosequencing (*Wilcoxon rank-sum test *p*-value < 0.05). **C.** Strip-plot showing the concordance of methylation levels between pooled and individual samples. Each dot represents the methylation level of one sample. **D.** Scatterplot shows the positive significant correlation between methylation in pooled and individual cfDNA samples for the 26 candidate CpG sites.

### Evaluation of candidate methylation biomarkers and further selection in individual samples

To evaluate the performance of the 26 candidate biomarkers in individual serum samples, methylation of the CpG positions of interest was quantified by bisulfite pyrosequencing in an independent cohort of 153 individuals (Table 1), which was randomly split between a biomarker evaluation cohort (30%, n=48) and a biomarker validation cohort (70%, n=105). Methylation levels of the v2 promoter region of the *SEPT9* gene were also quantified in all samples. A detailed description of the pyrosequenced regions and the number of CpG positions analyzed within each biomarker can be found in Supplementary Table 3. The construction of methylation models for CRC screening was conducted in two phases.

First, the 26 candidate biomarkers were evaluated in a subset of 48 serum samples (30%). The methylation levels of the 26 candidate biomarkers in both pooled (MethylationEPIC-derived) and individual cfDNA samples (bisulfite pyrosequencing) are shown in Figures 4A and B, respectively. There was a significant positive correlation (Pearson’s r > 0.6, *p*-value < 0.001) between cfDNA methylation in pooled and individual serum samples (Fig 4C, D). The performance (ROC curves, AUC, sensitivity, and specificity) of the 26 biomarkers for the detection of AN in the 48 samples of the biomarker evaluation cohort is presented in Supplementary Figure 1.

To further reduce the number of amplicons for validation, penalized logistic regressions LASSO and Elastic net were fitted to the complete list of 26 candidate biomarkers. Classification performance parameters were derived by leave-one-out cross-validation. Although only CG3, CG8, CG15, CG16, CG20, CG24, and CG25 reported statistically significant methylation differences (Wilcoxon rank-sum rest *p*-value < 0.05) between NN and AN (Fig. 4B), variable selection was applied to the whole set of 26 candidate biomarkers since it has been reported that prediction power is not always increased with variables significantly correlated with the outcome [42]. We produced sparse models containing two (CG3-*GALNT9* and CG15-*UPF3A*; AUC: 0.905, 95% CI 0.801-1) and four (CG3-*GALNT9*, CG15-*UPF3A*, CG5-*WARS,* and CG24-*LDB2*; AUC: 0.827, 95% CI 0.651-1) methylation biomarkers, derived by LASSO and Elastic net regularization, respectively. Also, the application of Elastic net to the 26 biomarkers, adding the clinical variables age and sex, generated a model containing 20 biomarkers and sex (AUC: 0.820, 95% CI 0.675-0.966). The four biomarkers selected by the sparse models are included in the 20-biomarker signature. Hence, this 20 biomarker set was selected to proceed with the validation phase. A detailed description of the biomarkers selected for validation is shown in Table 2, including the genomic region and the number of CpG sites analyzed by pyrosequencing.

**Table 2.**
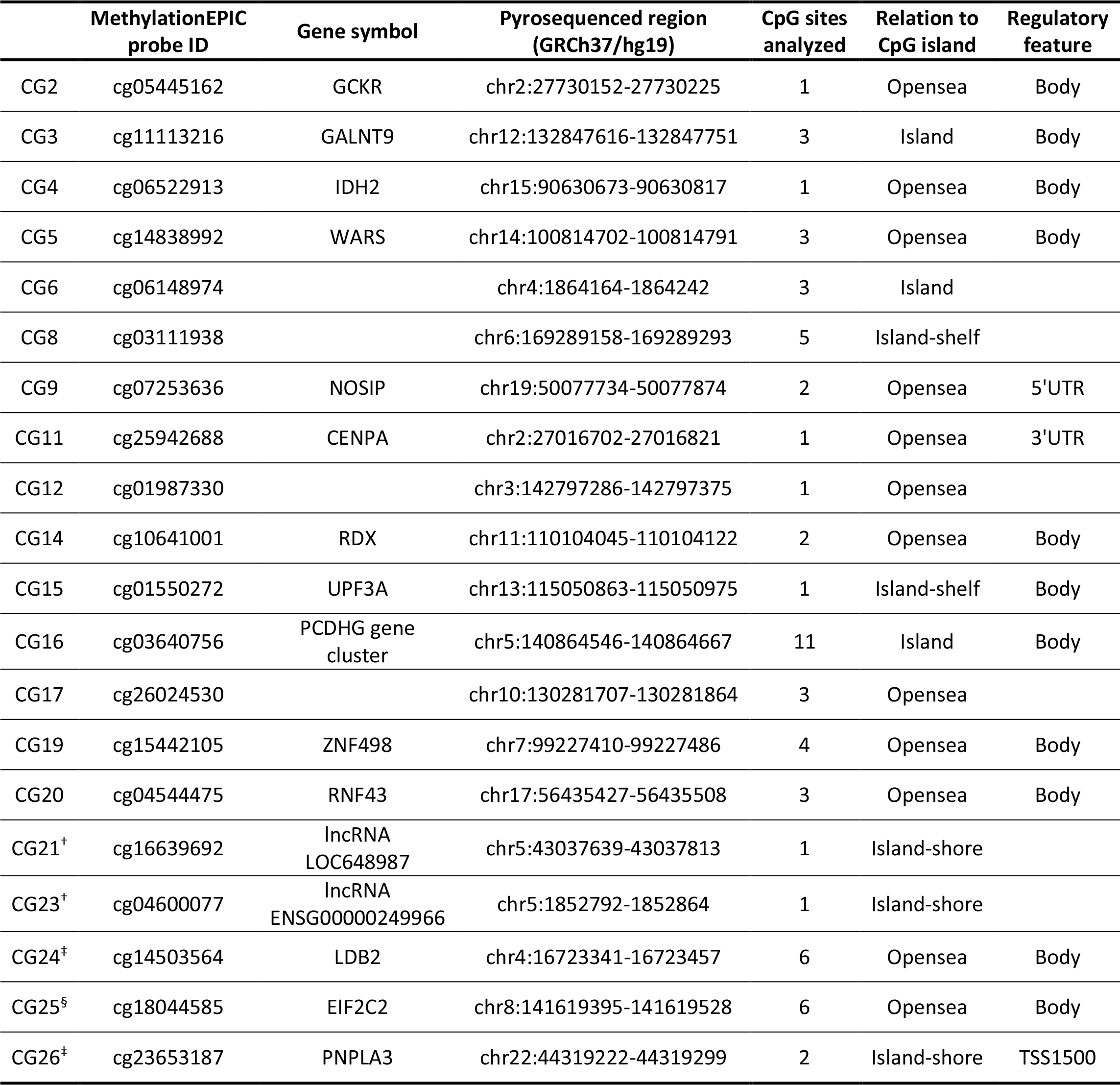
20 biomarkers selected for the validation phase. Regulatory features and relation to CpG island of biomarkers annotated according to the Methylation EPIC Manifest: CpG island: region of at least 200 bp with a CG content > 50% and an observed-to-expected CpG ratio ≥ 0.6; Island-shore: sequences 2 kb flanking the CpG island; Island-shelf: sequences 2 kb flanking shore regions; Opensea: sequences located outside these regions; Body: gene body (intragenic region); TSS1500: 1500 bp upstream the transcription start site. †Candidate biomarkers derived from the comparison with the external RRBS dataset; ‡candidate biomarkers derived from the NN vs P-AA classification; §candidate biomarker derived from the NN vs D-AA classification.

|  | MethylationEPIC probe ID | Gene symbol | Pyrosequenced region (GRCh37/hg19) | CpG sites analyzed | Relation to CpG island | Regulatory feature |
| --- | --- | --- | --- | --- | --- | --- |
| CG2 | cg05445162 | GCKR | chr2:27730152-27730225 | 1 | Opensea | Body |
| CG3 | cg11113216 | GALNT9 | chr12:132847616-132847751 | 3 | Island | Body |
| CG4 | cg06522913 | IDH2 | chr15:90630673-90630817 | 1 | Opensea | Body |
| CG5 | cg14838992 | WARS | chr14:100814702-100814791 | 3 | Opensea | Body |
| CG6 | cg06148974 |  | chr4:1864164-1864242 | 3 | Island |  |
| CG8 | cg03111938 |  | chr6:169289158-169289293 | 5 | Island-shelf |  |
| CG9 | cg07253636 | NOSIP | chr19:50077734-50077874 | 2 | Opensea | 5'UTR |
| CG11 | cg25942688 | CENPA | chr2:27016702-27016821 | 1 | Opensea | 3'UTR |
| CG12 | cg01987330 |  | chr3:142797286-142797375 | 1 | Opensea |  |
| CG14 | cg10641001 | RDX | chr11:110104045-110104122 | 2 | Opensea | Body |
| CG15 | cg01550272 | UPF3A | chr13:115050863-115050975 | 1 | Island-shelf | Body |
| CG16 | cg03640756 | PCDHG gene cluster | chr5:140864546-140864667 | 11 | Island | Body |
| CG17 | cg26024530 |  | chr10:130281707-130281864 | 3 | Opensea |  |
| CG19 | cg15442105 | ZNF498 | chr7:99227410-99227486 | 4 | Opensea | Body |
| CG20 | cg04544475 | RNF43 | chr17:56435427-56435508 | 3 | Opensea | Body |
| CG21<sup>†</sup> | cg16639692 | lncRNA LOC648987 | chr5:43037639-43037813 | 1 | Island-shore |  |
| CG23<sup>†</sup> | cg04600077 | lncRNA ENSG00000249966 | chr5:1852792-1852864 | 1 | Island-shore |  |
| CG24<sup>‡</sup> | cg14503564 | LDB2 | chr4:16723341-16723457 | 6 | Opensea | Body |
| CG25<sup>§</sup> | cg18044585 | EIF2C2 | chr8:141619395-141619528 | 6 | Opensea | Body |
| CG26<sup>‡</sup> | cg23653187 | PNPLA3 | chr22:44319222-44319299 | 2 | Island-shore | TSS1500 |

### Validation of the selected biomarkers and final model construction

The final 20 selected methylation biomarkers were then quantified in the remaining 105 serum samples (70%) (Fig. 5A) to validate their use as non-invasive biomarkers in individual samples. The performance (ROC curves, AUC, sensitivity, and specificity) of the 20 selected biomarkers for the detection of AN in the 105 samples (validation cohort) is presented in Supplementary Figure 2. Multivariate logistic regressions were fitted to the selected biomarker subsets obtained from the three best performing models from the previous step, to derive three new models containing 2 (CG3-*GALNT9* and CG15-*UPF3A*), 4 (CG3-*GALNT9*, CG15- *UPF3A*, CG5-*WARS*, and CG24-*LDB2*), and 20 biomarkers. Performances of the diagnostic prediction models in the biomarker validation cohort are summarized in Table 3, while ROC curves are provided in Figure 5B. ROC curves and performance parameters were obtained by leave-one-out cross-validation.

**Figure 5.**
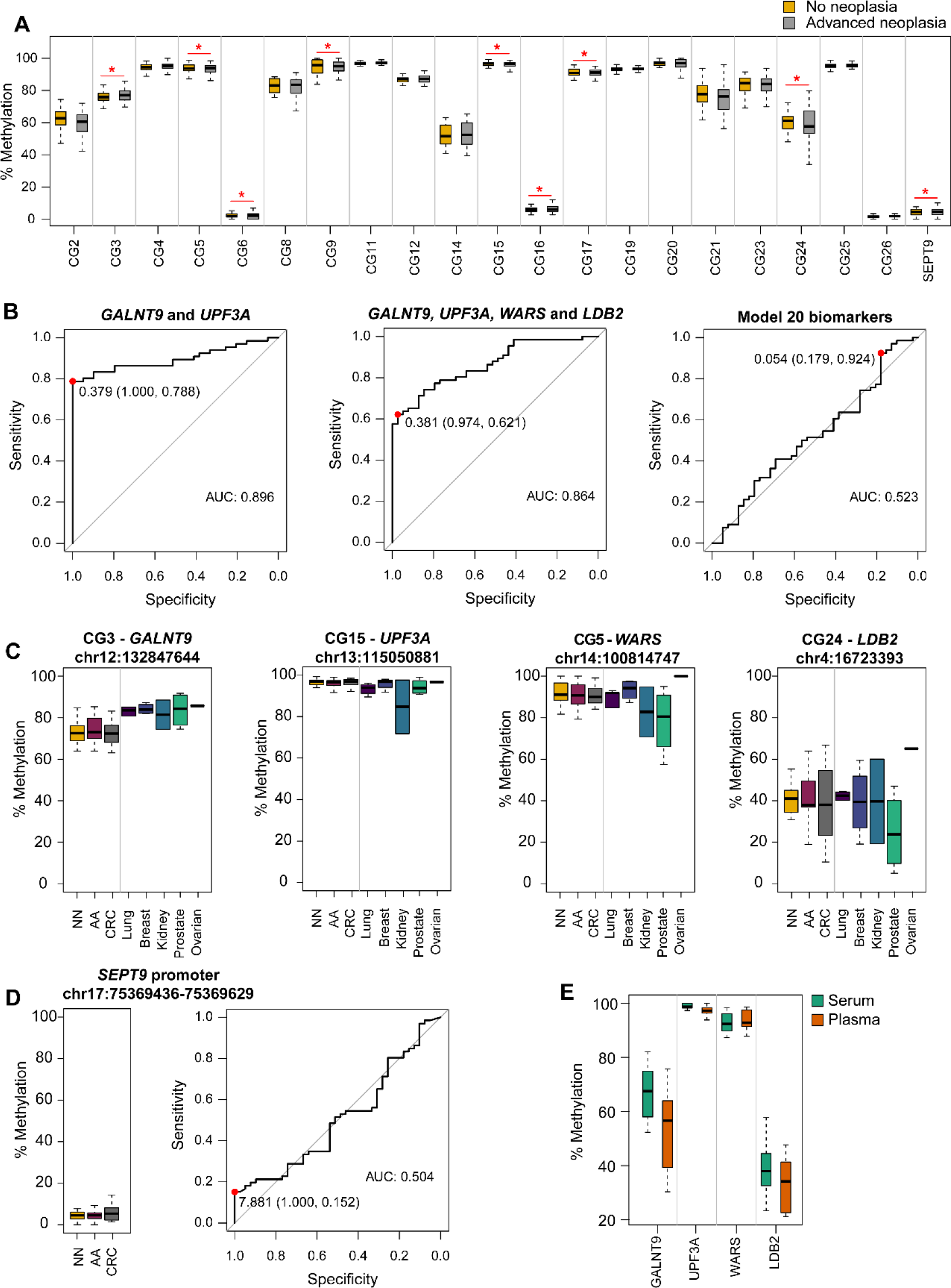
Diagnostic performance of the models and methylation levels in the biomarker validation cohort (n=105). **A.** Methylation levels of the final 20 selected biomarkers in the validation cohort (*Wilcoxon rank-sum test *p*-value < 0.05). B. ROC curve analysis and AUC for the three models evaluated for CRC screening, derived by leave-one-out cross- validation (*GALNT9*: CG3; *UPF3A*: CG15; *WARS*: CG5; *LDB2*: CG24). The red dots indicate the sensitivity and specificity values for the best cut-offs based on the Youden Index method. **C.** Serum methylation levels of CG3-*GALNT9*, CG15- *UPF3A*, CG5-*WARS*, and CG24-*LDB2* in the biomarker validation cohort (n=105), and in lung, breast, kidney, prostate, and ovarian cancer cases (n=16). **D.** Methylation levels and classification performance (ROC curve) of the SEPT9 promoter. The red dot indicates the best sensitivity and specificity values (Youden Index). **E.** Comparison of methylation levels between matched serum and plasma samples. AUC: area under the curve; NN: no neoplasia; AA: advanced adenomas; CRC: colorectal cancer.

**Table 3.** Performance of the models and *SEPT9* for advanced neoplasia detection in the biomarker validation cohort. The biomarker validation cohort includes 105 serum samples. AA and CRC detection rates are also shown. No significant differences were found between the detection of distal versus proximal AA (Fisher’s exact test p-value>0.05). ROC curves and performance parameters were derived by the leave-one-out cross-validation approach. AUC: area under the curve; AA: advanced adenoma; CRC: colorectal cancer; NPV: negative predictive value; PPV: positive predictive value; *GALNT9*: CG3; *UPF3A*: CG15; *WARS*: CG5; *LDB2*: CG24.

|  | Advanced Neoplasia classification |  |  |  |  | AA detection % |  |  | CRC detection % |  |  |  |  |
| --- | --- | --- | --- | --- | --- | --- | --- | --- | --- | --- | --- | --- | --- |
|  | AUC<br>(95% CI) | Specificity %<br>(95% CI) | Sensitivity %<br>(95% CI) | NPV %<br>(95% CI) | PPV %<br>(95% CI) | D-AA | P-AA | All | Stage<br>I | Stage<br>II | Stage<br>III | Stage<br>IV | All |
| ***GALNT9, UPF3A*** | 0.896<br>(0.835–0.958) | 100<br>(91–100) | 78.8<br>(67–88) | 73.6<br>(60–85) | 100<br>(93–100) | 87 | 68.4 | 78.6 | 87.5 | 100 | 100 | 33.3 | 79.2 |
| ***GALNT9, UPF3A, WARS, LDB2*** | 0.864<br>(0.798–0.931) | 97.4<br>(87–100) | 62.1<br>(49–74) | 60.3<br>(47–72) | 97.6<br>(87–100) | 78.3 | 52.6 | 66.7 | 37.5 | 75 | 100 | 33.3 | 54.2 |
| **20 methylation biomarkers, sex** | 0.553<br>(0.407–0.639) | 17.9<br>(8–34) | 92.4<br>(83–97) | 58.3<br>(28–85) | 65.6<br>(55–75) | 91.3 | 94.7 | 92.9 | 87.5 | 100 | 100 | 83.3 | 91.7 |
| ***SEPT9*** | 0.504<br>(0.389–0.618) | 100<br>(91–100) | 15.1<br>(8–26) | 41.1<br>(31–52) | 100<br>(69–100) | 4.3 | 10.5 | 7.1 | 0 | 37.5 | 50 | 50 | 29.2 |

The model composed of *GALNT9* and *UPF3A* showed the best diagnostic performance for CRC screening, yielding an AUC of 0.896 (95% CI 0.835-0.958), discriminating advanced neoplasia from no neoplasia controls with 78.8% sensitivity and 100% specificity. This model identified 33 out of 42 AA cases (78.6%) and 19 out of 24 CRC patients (79.2%), with notable detection of early-stage CRC (87.5% and 100% for stages I and II, respectively). This 2-biomarker panel detected 87% of distal AA and 68.4% proximal AA. On the other hand, the resulting AUC for the model containing *GALNT9*, *UPF3A*, *WARS*, and *LDB2* was 0.864 (95% CI 0.798- 0.931), with a sensitivity of 62.1% and a specificity of 97.4%, detecting 28 out of 42 AA (66.7%; 78.3% distal and 52.6% proximal), and 13 out of 24 CRC cases (54.2%; 37.5 stage I and 75% stage II). These two models demonstrated no significant differences in the detection of distal AA compared to proximal ones (Fisher’s exact test *p*-value > 0.05). Finally, the model containing 20 biomarkers and sex, reported the highest sensitivity (92.4%) and the highest detection rate for AA and CRC (92.9% and 91.7%, respectively), but with considerably less specificity (17.9%) when compared to the previous models. The logistic classification rules of the model containing *GALNT9*, *UPF3A*, *WARS*, and *LDB2* and the *GANL9*/*UPF3A* model are detailed in the supplementary information (see Additional File 2). *GALNT9* (CG3) was hypermethylated in AN, whilst *UPF3A* (CG15), *WARS* (CG5), and *LDB2* (CG24) showed hypomethylation in AN. Differences in the methylation levels between NN and AN of these biomarkers were statistically significant (Wilcoxon rank-sum test *p*-value < 0.05; Fig. 5A).

Since plasma samples are most commonly used as a source of cfDNA, methylation of *GALNT9*, *UPF3A*, *WARS*, and *LDB2* was also evaluated in 8 pairs of matched serum and plasma samples. No statistical differences were found between serum and plasma methylation levels (Wilcoxon signed-rank test *p*-value > 0.01) (Fig. 5E).

### Analysis and performance of Septin9 methylation

The methylation EPIC array contains three probes targeting the region of the v2 promoter of the *SEPT9* gene commonly evaluated for CRC screening [43, 44] (cg02884239, cg20275528, cg12783819). None of them reported significance for the detection of AN in the epigenome-wide differential methylation analysis performed on the 28 cfDNA pooled samples, and showed an average of 0.14% methylation difference between NN and AN (Fig. 4A). We reported hypomethylation of *SEPT9* in AN in both the biomarker evaluation cohort (n=48) (Fig. 4B) and the biomarker validation cohort (n=105) (Fig. 5A). The diagnostic performance of the *SEPT9* promoter methylation was also evaluated in the biomarker validation cohort, with an AUC of 0.504 (95% CI 0.389-0.618) for AN detection and sensitivity and specificity values of 15.2% and 100%, respectively (Table 3, Figure 5D). Only 7.1% of AA and 29.2% of CRC were detected.

### Evaluation of the classification models in non-colorectal tumors

To assess the ability of the models to specifically detect colorectal advanced adenomas and cancer, methylation of *GALNT9*, *UPF3A*, *WARS,* and *LDB2* was quantified in serum samples from patients with lung, breast, kidney, prostate, and ovarian cancer (n=16) (Fig. 5C, Supplementary Table 1). When applied to different cancer types, the *GALNT9*/*UPF3A* model misclassified as advanced colorectal neoplasia 3 out of 16 cancer cases (18.75%: prostate, kidney, and breast cancer). On the other hand, the model composed of *GALNT9*, *UPF3A*, *WARS*, and *LDB2* incorrectly identified 1 out of 16 cancer cases (6.25%: ovarian cancer) as advanced neoplasia.

## DISCUSSION

Early detection has proven to be the most effective strategy to reduce both the incidence and mortality of CRC [6, 7]. The FIT is the most widely implemented and recommended non-invasive test for CRC screening despite having a modest sensitivity for the detection of premalignant AA (22-56%, 90-94% specificity) [9–12]. Also, most current screening programs based on FIT followed by a confirmatory colonoscopy suffer from low participation rates (43.8% worldwide) [4, 8]. Currently, there is no non-invasive biomarker for the early detection of CRC and AA that meets all the characteristics of an ideal screening test: easy to perform, safe, highly specific and sensitive, cost-effective, and well accepted by patients. To this end, the development of liquid biopsy technology has shown to be a promising approach for CRC screening, diagnosis, follow-up, and treatment guidance [15], with the potential to be integrated into the clinic.

In this study, we first conducted an epigenome-wide analysis with the MethylationEPIC array using a cfDNA pooling approach to discover potential blood-based biomarkers for the joint detection of AA and CRC. The selection process of the candidate biomarkers derived from the epigenome-wide analysis was conducted by penalized logistic regression (LASSO and Elastic net regularizations). After prioritization of candidate biomarkers and evaluating their methylation levels in individual samples from an independent cohort, we developed and cross-validated three prediction models for the detection of AN (AA and CRC). The first one, the 20 methylation biomarkers with sex, yielded a sensitivity of 92.4% for AN, at 18% specificity. Despite its high sensitivity and highest detection rate for AA (92.9%) and CRC (91.7%), such low specificity is not cost-effective for screening programs. Secondly, the model composed of *GALNT9*, *UPF3A*, *WARS,* and *LDB2* reported 62.1% sensitivity and 97.4% specificity, whilst the *GALNT9*/*UPF3A* model discriminated AN with 78.8% sensitivity and 100% specificity, showing the best prediction performance for CRC screening. The sensitivities reported for our biomarkers are comparable to that of FIT for CRC detection (73-88%) and higher than its sensitivity for AA (22-56%) [12]. The combination *GALNT9*/*UPF3A* also fulfills the main objective of CRC screening, that is the detection of preclinical CRC and premalignant AA, as reported suitable detection rate for AA (78.6%) and early CRC stages I and II (87.5% and 100%, respectively). Also, no statistically significant differences were reported between the detection of distal (87%) and proximal (68.4%) AA, in contrast with the FIT that performs better for distal lesions than proximal ones [45]. The decreased detection of stage IV tumors (33.3%) would not affect the effectiveness of CRC screening, since individuals with late-stage cancers are commonly symptomatic and therefore will not be enrolled in screening programs targeting the average-risk population.

Among the four methylation biomarkers, only *UPF3A* has been previously related to CRC. Located in a CpG island shelf, we reported the hypomethylation of *UPF3A* in AN; and high expression levels of this gene were associated with TNM stage, liver metastasis, and recurrence in CRC [46]. *GALNT9* is also located in a CpG island and showed hypermethylation in AN, which was also reported in brain metastasis from primary breast cancer [47]. On the other hand, *WARS* and *LDB2* also show hypomethylation in AN but are located within opensea regions. The methylation of regions outside GpG islands does not always directly relate to gene silencing. High expression levels of *WARS* were found in high microsatellite-instable gastrointestinal adenocarcinomas, associated with poor prognosis [48], whilst a decreased expression of *LDB2* was associated with a more favorable outcome in lung adenocarcinoma patients [49].

Nowadays, the only non-invasive methylation biomarker approved by the FDA for the detection of CRC in blood is based on the methylation of the SEPT9 promoter [50]. Nevertheless, the results on the diagnostic performance of this plasma biomarker are variable and inconsistent, with sensitivities ranging from 48.2- 95.6% for CRC and 11.2-35% for AA, with 79.1-99.1% specificity. In an asymptomatic average-risk cohort, SEPT9 showed lower screening performance than FIT (sensitivity: 68% vs. 79%; specificity: 80% vs. 94%, respectively), reviewed in [51–54]. In our study, we evaluated the performance of SEPT9 in both our discovery and validation cohorts. The 3 CpG sites targeting SEPT9 interrogated in the MethylationEPIC BeadChip were not differentially methylated between NN and AN cfDNA pooled samples, and in the final validation cohort, the sensitivity for AN and AA resulted in 15.1% and 7%, respectively, with 100% specificity (pyrosequencing 5 CpG positions). Unlike the commercial test, based on qPCR positive reactions in plasma cfDNA [55], we quantified the methylation of the SEPT9 promoter by pyrosequencing in serum samples and thus the results may not be fully comparable.

Several blood-based methylation biomarkers have recently emerged for the early detection of CRC. Plasma BCAT1 and IKZF1 methylation reported sensitivities of 66% and 5% for CRC and AA, respectively, at 95% specificity [56]. Methylation markers C9orf50, KCNQ5, and CLIP4 reported 85% sensitivity and 99% specificity for CRC detection [57]. Methylated SFRP2 and SDC2 detected CRC and AA with sensitivities of 76.2% and 58.3%, and 87.9% specificity [58]. A single plasma methylation biomarker, cg10673833, demonstrated 89.7% sensitivity and 86.8% specificity for CRC, but the sensitivity dropped up to 33.3% for precancerous lesions [59].

In contrast with the aforementioned available studies, based on results reported in cell lines, tissue, or stool, the design of our study targets the final sample format for both the discovery and validation phases, enhancing the possibility to discover and translate robust non-invasive biomarkers. From our proposed models, the combination *GALNT9*/*UPF3A* showed a convenient 78.8% sensitivity for the joint detection of AA and CRC (detecting 78.6% AA and 79.2% CRC: 87.5% stage I and 100% stage II), and an advantageous specificity of 100%, superior to other reported potential biomarkers. A cost-effective biomarker for CRC screening should present a low false-positive rate to minimize the number of unnecessary, invasive, and expensive colonoscopies. Thus, the highest specificity is sought for CRC screening.

In the ongoing validation of CRC screening biomarkers, it is important to consider data on protocol acceptability. A blood-based test has the potential to improve compliance and participation in CRC screening, as reported by a randomized controlled trial where 99.5% of subjects who were offered a SEPT9 as the first option for CRC screening completed the test within six weeks, compared to 88.1% of participants in the FIT arm [60]. It was also reported that 83% of individuals enrolled for screening who refused colonoscopy, preferred a blood-based test [14]. To optimize the participation in CRC screening, perhaps both a fecal and a blood-based test should be offered to target different preferences. A blood test could be proposed as a screening option to invitees refusing FIT and other guidelines recommended tests. This is indeed the indication for the SEPT9 test in average-risk individuals older than 50 [54, 61]. Another option for implementing a blood test in screening programs is triaging FIT-positive individuals for improved selection to colonoscopy [62]. Figure 6 shows a schematic representation of the possible implementation of a blood test in CRC screening, both as an alternative to FIT aiming to increase participation rates, and as a triage approach to optimize selection to colonoscopy.

**Figure 6.**
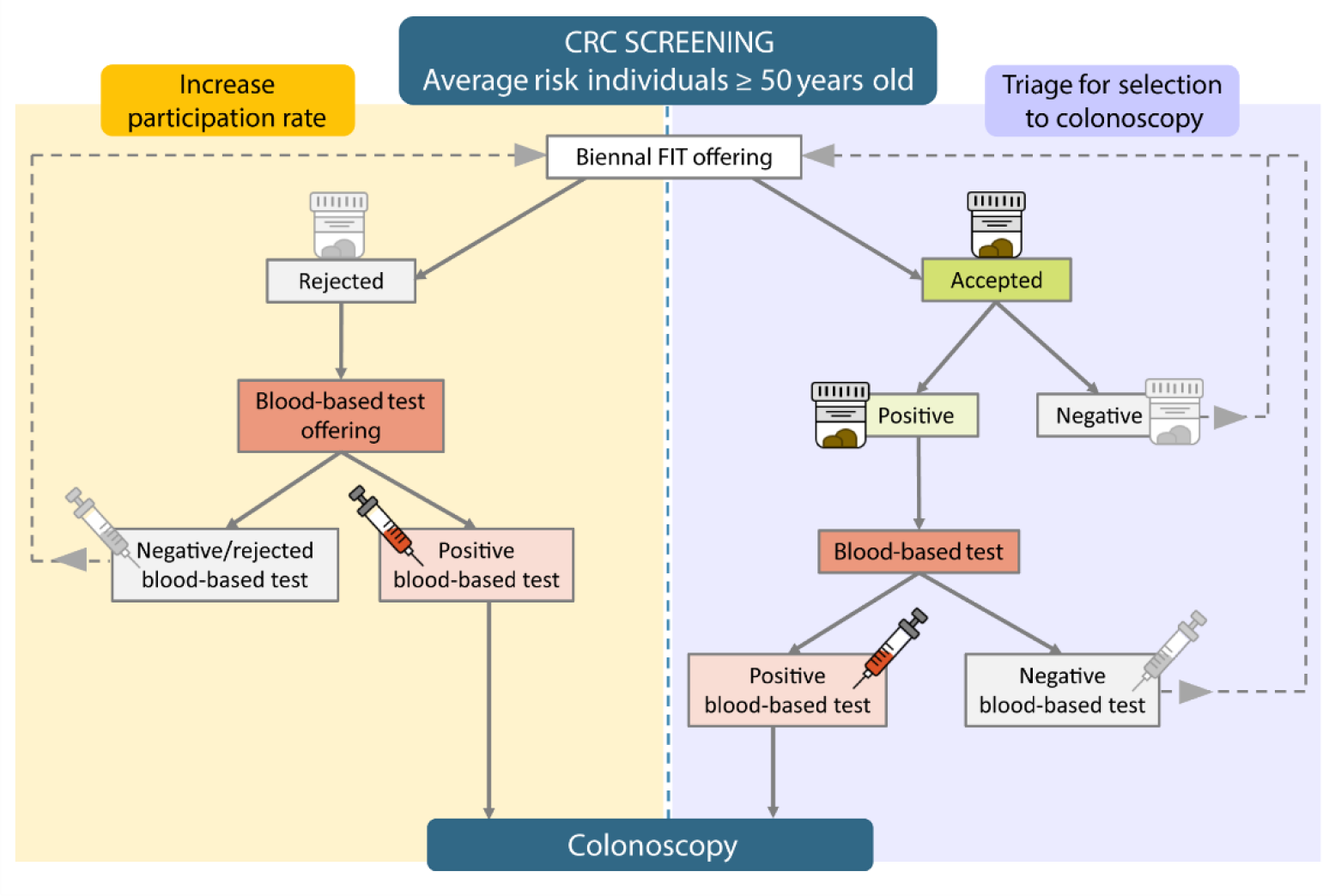
Schematic representation of the potential implementation of a blood-based test in CRC screening. To target different sample preferences and improve participation rates, a blood test could be offered to individuals refusing the FIT (left arm). In combination with the FIT, the blood test may improve selection to follow-up colonoscopy after a positive FIT (right arm). A FIT test would be offered every two years to individuals rejecting both the FIT and the blood test, and to individuals with a previous negative result in either the FIT or blood-based test.

To the best of our knowledge, this is the first study conducting a serum-based discovery and validation of cfDNA methylation biomarkers for CRC screening. The reported results underline the feasibility of cfDNA pooled samples as an affordable approach for biomarker discovery. Sample pooling strategies also allow increasing the DNA input when small amounts are available [21,22,63,64]. Our multicentre cohort includes not only CRC, AA, and healthy controls, but also benign pathologies typically found during screening programs that influence the specificity, such as non-advanced adenomas, hemorrhoids, and diverticula. It is worth mentioning that our healthy individuals are not *self-declared* but colonoscopically confirmed and that the criteria of inclusion and age range of the patients followed the USPSTF guidelines for CRC screening [39].

Also, we have checked that there is no difference in the methylation levels of our biomarkers between serum and plasma cfDNA, and the low volume of serum used (up to 2 mL) is feasible in clinical practice. The ability of our methylation biomarkers to specifically detect CRC was also assessed, incorrectly identifying only 18.75% of other-cancer samples as colorectal advanced neoplasia by the *GALNT9*/*UPF3A* model, and 6.25% by the *GALNT9*, *UPF3A*, *WARS*, and *LDB2* model.

Nevertheless, our study has some limitations. Firstly, CRC cases were mostly diagnosed having symptoms, and secondly, the proportions of CRC cases, tumor stages, and the rest of pathologies are not fully representative of a screening population. A large prospective validation in an asymptomatic average-risk screening population will be necessary before the implementation of any of the proposed methylation models.

## CONCLUSIONS

We have discovered and reported *GALNT9*, *UPF3A*, *WARS*, and *LDB2* as new non-invasive biomarkers for the early detection of colorectal cancer and advanced adenomas, regardless of the lesion location. We propose that the combination of methylated *GALNT9*/*UPF3A* is the most promising to serve as a highly specific blood-based test for screening and detection of CRC at an early and curable stage, even at the premalignant lesion phase. Our results need further validation in a real screening setting.

## ABBREVIATIONS

AA: advanced adenoma; AN: advanced neoplasia; AUC: area under the curve; BEN: benign pathologies (hemorrhoids and diverticula); cfDNA: circulating cell-free DNA; CRC: colorectal cancer; D-AA: individuals with advanced adenomas of both distal and proximal locations; DMP: Differentially methylated position; FDR: false discovery rate; FIT: fecal immunochemical test; NCF: no colorectal findings; NN: no neoplasia; NPV: negative predictive value; PPV: positive predictive value; ROC: Receiver-operating characteristic.

## DECLARATIONS

### Ethics approval and consent to participate

Written informed consent was obtained from all patients with approval by the Ethics Committee for Clinical Research of Galicia (2018/008). The study was conducted according to the clinical and ethical principles of the Spanish Government and the Declaration of Helsinki.

### Availability of data and materials

The Infinium MethylationEPIC data from all the pooled samples generated and analyzed during this study have been deposited in the NCBI Gene Expression Omnibus (GEO) (www.ncbi.nlm.nih.gov/geo) and are accessible through GEO Series accession number GSE186381

[https://www.ncbi.nlm.nih.gov/geo/query/acc.cgi?acc=GSE186381]

### Author’ contributions

LD conceived and designed the study. LD, DGC, JC, and ME supervised the study. MGG, LD, and SM contributed to the sample preparation and data acquisition. MGG, NP, and MRG performed the analysis and interpretation of data. MGG, MRG, NP, DGC, and LD contributed to the experimental design. JC, LB, AE, AC, FB, and RJ provided clinical advice for the study design, collection, and management of clinical data. MGG and LD prepared the manuscript. All authors critically reviewed and approved the final manuscript.

## Supporting information

Supplementary information about bioinformatics preprocessing of microarray data, primers and PCR conditions, and final classification rules

## Data Availability

https://www.ncbi.nlm.nih.gov/geo/query/acc.cgi?acc=GSE186381

## Acknowledgments

We would like to thank Dr. Vicenta Soledad Martínez Zorzano, Dr. Francisco Javier Rodríguez Berrocal and Dr. María Páez de la Cadena for their support and scientific advice. Also, we would like to thank Dr. Juan Miguel Gómez Zumaquero for the pyrosequencing support. The samples and clinical data of patients included in this study were provided by: the HGUA Biobank (PT13 / 0010/0044), integrated into the *Red Nacional de Biobancos* and in the *Red Valenciana de Biobancos; SERGAS Biobanco A Coruña*; Basque Biobank/Biodonostia Node; Biobank at the Galicia Sur Health Research Institute; Biobank of Hospital Clínic, Barcelona – IDIBAPS; and CHUS’s Biobank (University Hospital Complex of Santiago de Compostela) with the approval of the respective Ethical and Scientific Committees, and have been processed following standard procedures.

## SUPPLEMENTARY INFORMATION

**Additional File 1: Supplementary Table 1.** Epidemiological and clinical data of patients with other tumors (n=16). **Supplementary Table 2.** Description of cfDNA pooled samples. **Supplementary Table 3.** Primers, PCR conditions, and amplicon details for biomarker evaluation by pyrosequencing. **Supplementary Table 4.** Description of the CpG candidate biomarkers obtained after the epigenome-wide methylation analysis.

**Additional File 2:** Details about the bioinformatics preprocessing of methylation microarray data for biomarker discovery. PCR conditions and primers for biomarker evaluation and validation in individual serum samples. Decision rules derived from the final models for de detection of colorectal advanced neoplasia.

**Additional File 3: Supplementary Figure 1**. ROC curve analysis for the 26 candidate biomarkers and *SEPT9* for NN versus AN classification in the biomarker evaluation cohort (n=48). **Supplementary Figure 2.** ROC curve analysis for the 20 selected biomarkers and *SEPT9* for NN versus AN classification in the biomarker validation cohort (n=105).

