## Supplementary Figure 1; Supplementary Figure 2 for "Serum methylation of *GALNT9*, *UPF3A*, *WARS,* and *LDB2* as non-invasive biomarkers for the early detection of colorectal cancer and premalignant adenomas"

Supplementary figure 1. ROC curve analysis for the 26 candidate biomarkers and SEPT9 for NN versus AN classification in the biomarker evaluation cohort (n=48). The red dots indicate the best cut−offs based on the Youden Index method. AN: advanced neoplasia; AUC: area under the curve; NN: no neoplasia; Se: sensitivity; Sp: specificity; YI: Youden Index.

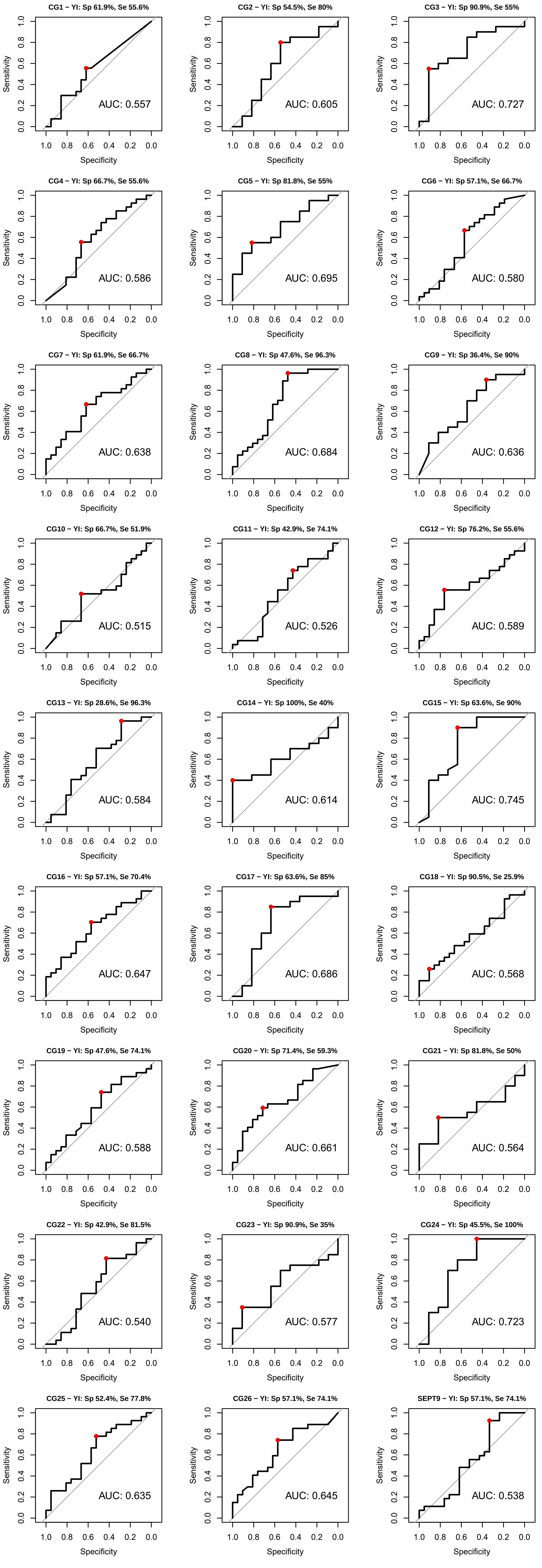

Supplementary figure 2. ROC curve analysis for the 20 selected biomarkers and SEPT9 for NN versus AN classification in the biomarker validation cohort (n=105). The red dots indicate the best cut−offs based on the Youden Index method. AN: advanced neoplasia; AUC: area under the curve; NN: no neoplasia; Se: sensitivity; Sp: specificity; YI: Youden Index.

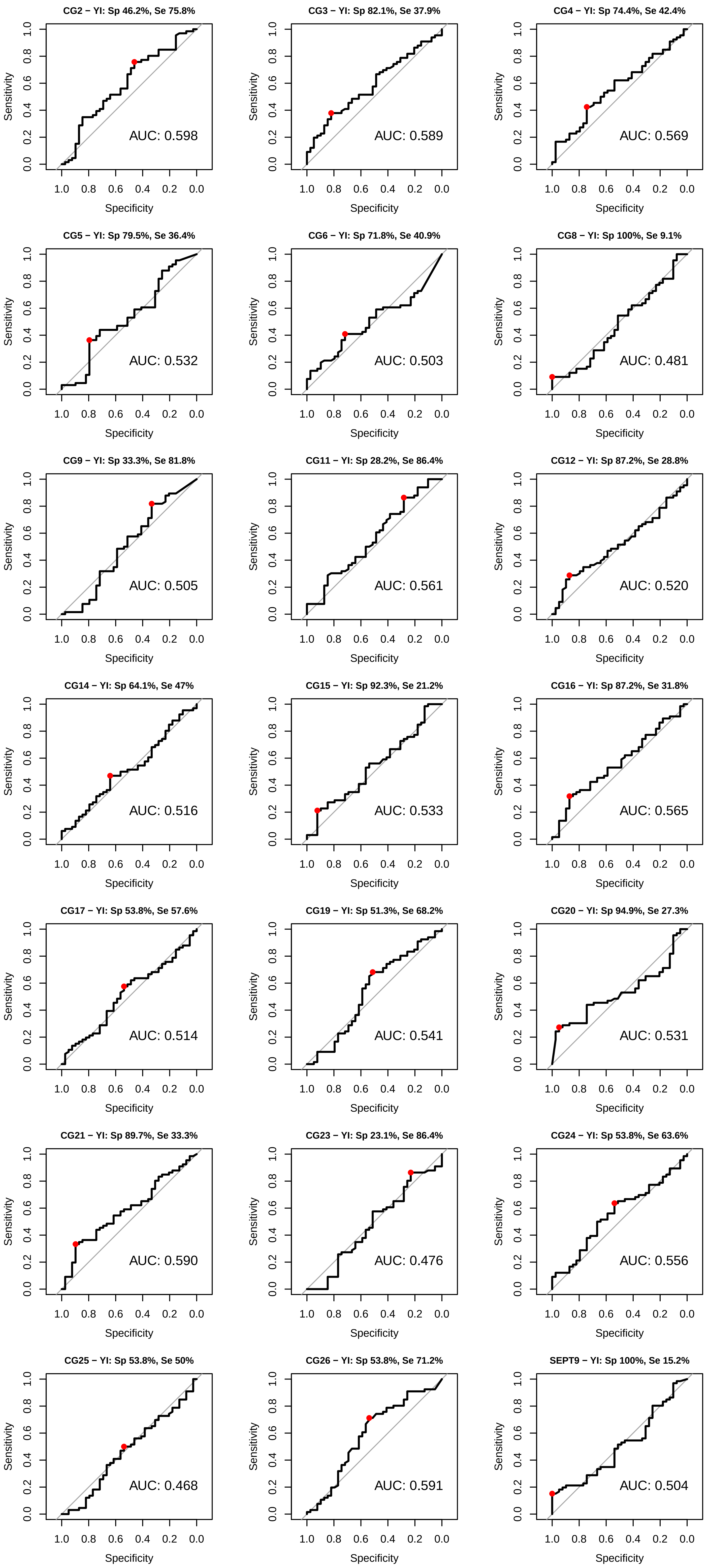
