## Supplementary Table 1; Supplementary Table 2; Supplementary Table 3; Supplementary Table 4 for "Serum methylation of *GALNT9*, *UPF3A*, *WARS,* and *LDB2* as non-invasive biomarkers for the early detection of colorectal cancer and premalignant adenomas"

**Supplementary Table 1.** Epidemiological and clinical data of patients with other tumors (n=16).

| | |
|---|---|
| **Age median (range)** | 61 (35-79) |
| **Male** | 8 |
| **Female** | 8 |
| **Breast cancer** | **4** |
| Luminal A, Luminal B | 1 |
| Luminal B, HER2-negative | 3 |
| **Kidney cancer** | **2** |
| Papillary transitional cell carcinoma | 2 |
| **Lung cancer** | **5** |
| Adenocarcinoma | 2 |
| Squamous cell carcinoma | 2 |
| Small cell neuroendocrine carcinoma | 1 |
| **Prostate cancer** | **4** |
| Adenocarcinoma | 3 |
| Acinar adenocarcinoma | 1 |
| **Ovarian cancer** | **1** |
| High grade serous carcinoma | 1 |

**Supplementary table 2. Description of cfDNA pooled samples**.

| Pool type | Age median (range) | Total amount DNA (ng) | Pathology description |
|---|---|---|---|
| NCF | 63 (52-71) | 160.0 | Each pool contained 10 individuals with no colorectal findings |
| | 62 (53-71) | 179.2 | |
| | 60.5 (54-71) | 190.4 | |
| BEN | 62 (53-71) | 142.6 | Each pool contained 5 individuals with hemorrhoids and 5 individuals with diverticula. |
| | 61 (51-70) | 164.8 | |
| | 62.5 (52-72) | 210.2 | |
| | 61.5 (52-70) | 206.1 | |
| | 62.5 (52-72) | 403.2 | |
| NAA | 62 (52-72) | 225.6 | Each pool contained 10 individuals with colorectal tubular adenomas without high-grade dysplasia and less than 10 mm in size. |
| | 61 (52-71) | 176.0 | |
| | 61.5 (52-72) | 195.2 | |
| | 62 (51-72) | 214.4 | |
| | 61.5 (51-72) | 169.6 | |
| D-AA | 61.5 (53-72) | 156.6 | Each pool contains 10 individuals with adenomas greater than 10 mm in size, and/or with villous histological component, and/or high-grade dysplasia. The most severe lesion was located in the distal colon, but other adenomas may be present also in the proximal colon. |
| | 62 (51-71) | 184.0 | |
| | 62 (52-70) | 203.2 | |
| | 62 (54-70) | 336.0 | |
| | 63 (54-69) | 225.6 | |
| P-AA | 60 (52-72) | 332.8 | Each pool contains 10 individuals with adenomas greater than 10 mm in size, and/or with villous histological component, and/or high-grade dysplasia. All advanced adenomas were of proximal location. |
| | 61.5 (52-69) | 256.0 | |
| | 62.5 (53-72) | 158.1 | |
| | 61 (56-70) | 128.2 | |
| | 61 (55-71) | 61.9 | |
| CRC I/II | 60.5 (53-71) | 182.4 | 6 CRC stage I and 4 CRC stage II |
| | 62 (51-71) | 185.6 | 5 CRC stage I and 5 CRC stage II |
| | 62 (51-70) | 169.6 | 6 CRC stage I and 4 CRC stage II |
| CRC III/IV | 62.5 (53-72) | 147.0 | 7 CRC stage III and 3 CRC stage IV |
| | 62.5 (52-71) | 157.5 | 6 CRC stage III and 4 CRC stage IV |

Pools were constructed with equal amounts of cfDNA from 5 men and 5 women from the same pathological group, recruitment hospital- and age-matched. NCF: no colorectal findings; BEN: benign pathology; NAA: non-advanced adenomas; D-AA: distal advanced adenomas; P-AA: proximal advanced adenomas; CRC: colorectal cancer.

**Supplementary table 3.** Primers, PCR conditions and amplicon details for biomarker evaluation by pyrosequencing. Biomarkers included in the same multiplex PCR reaction are grouped together. Biotin labelled primers are highlighted with italics.

| Biomarkers | Primers | Singleplex PCR temperature (°C) | Singleplex PCR cycles | Singleplex amplicon length (bp) | Pyrosequenced region (GRCh37/hg19) | Pyrosequenced region length (bp) | Amount of CpG sites analysed |
|---|---|---|---|---|---|---|---|
| CG2 | PCR→ GTGATATGTTTAATTAGAAGGTTGAGTTTA<br>PCR← *CACACTAATAATCTCCCCAACT*<br>Sequencing→ AATTAGAAGGTTGAGTTTATTAA | 60 | 25 | 85 | chr2:27730152-27730225 | 74 | 1 |
| CG3 | PCR→ TTTAAAAATTAAGTAGAGGGGAGAGTAGGT<br>PCR← *ACCCCACATAACCACTACTACC*<br>Sequencing→ AGGGGAGAGTAGGTG | 60 | 25 | 152 | chr12:132847616-132847751 | 136 | 3 |
| CG5 | PCR→ GGGAGAAGTATAATGTTGGGAGGTTTGTAA<br>PCR← *TCTCCAATACCCCCCAAAAC*<br>Sequencing→ TTTAGGTTGAGTAGAGGTA | 58 | 25 | 145 | chr14:100814702-100814791 | 90 | 3 |
| CG11 | PCR→ GGGGATTTTTTAGAGTTATGATTAGAT<br>PCR← *AATCATACAATCTTCTCCTTCTCA*<br>Sequencing→ GAGTTATGATTAGATTTAATGGA | 60 | 20 | 132 | chr2:27016702-27016821 | 120 | 1 |
| CG21 | PCR→ GTTTTGGGTTTTAGTAAGTTTTATAGAAGT<br>PCR← *ACTAACCCTCAACTTTATACTATCT*<br>Sequencing→ GGTTTTAGTAAGTTTTATAGAAGTA | 58 | 20 | 181 | chr5:43037639-43037813 | 175 | 1 |
| SEPT9 | PCR→ *AGGGGGTTTAGGGGTTTTT*<br>PCR← CCAACCCAACACCCACCT<br>Sequencing← AAATCCCAAATAATCCCATCC | 58 | 25 | 215 | chr17:75369436-75369629 | 194 | 5 |

**Supplementary table 3 (continuation).**

| Biomarkers | Primers | Singleplex PCR temperature (°C) | Singleplex PCR cycles | Singleplex amplicon length (bp) | Pyrosequenced region (GRCh37/hg19) | Pyrosequenced region length (bp) | Amount of CpG sites analysed |
|---|---|---|---|---|---|---|---|
| CG8 | PCR→ *GTGTGTTGATTGTGGATAGGT*<br>PCR← ACAACCATAAAATTCTACTAAATCTAAAC<br>Sequencing← AATTCTACTAAATCTAAACAATAAT | 58 | 20 | 146 | chr6:169289158-169289293 | 136 | 5 |
| CG9 | PCR→ GGGTTTGGATAGTTATAGGATGT<br>PCR← *TCCAACCTCAAAAACTAAAAAAAATAAATC*<br>Sequencing→ ATTGGAAAATATATAGTTGTAGT | 58 | 20 | 175 | chr19:50077734-50077874 | 141 | 2 |
| CG12 | PCR→ TTGTGATTGGTGGTTGTAGGT<br>PCR← *AACTTCCCTACCCTATATTAAAACCACTA*<br>Sequencing →GTGGTGAGAAGAAAATAATT | 58 | 25 | 109 | chr3:142797286-142797375 | 90 | 1 |
| CG13 | PCR→ AGATAGGGTTGTTTAGTTTTAATGATAATA<br>PCR← *ATCTCAAATCTACCCCTCTCAAAAATACAA*<br>Sequencing→ ATTATTGGTGTTTATTAGTTGA | 58 | 20 | 223 | chr1:28160760-28160884 | 125 | 2 |
| CG15 | PCR→ AGGGAGTATGTTATTTGTTATTGAATGA<br>PCR← *AATATTTTTATACCAACCTCCACTATC*<br>Sequencing→ TAGGTTTTGTGGGTG | 60 | 20 | 156 | chr13:115050863-115050975 | 113 | 1 |
| CG1 | PCR→ GGTTTTTGTAAATAGTTGTATTGAAGTAG<br>PCR← *AACTACTAAATAACACCAACAACATC*<br>Sequencing→ AAATAGGTATAAAGAAGATTGT | 60 | 20 | 129 | chr15:83563756-83563833 | 78 | 1 |
| CG4 | PCR→ GGGAAGGGAAGAAAGGTTATAGAGTAT<br>PCR← *CCCCCTACAATCCATCTCAAATTTTAC*<br>Sequencing→ GTTAAAGATTTTTTGGAAGATG | 60 | 20 | 190 | chr15:90630673-90630817 | 145 | 2 |
| CG18 | PCR→ ATTTGGAGGTTTTGTGTTTGT<br>PCR← *CTAACCCCCCTAAAACATCAAATAACAATC*<br>Sequencing→ TTTGTTGGTAGGGGTA | 60 | 25 | 104 | chr22:45094510-45094597 | 88 | 2 |
| CG20 | PCR→ TTGGGTTAGGTTTTTTGTTATGTTATT<br>PCR← *AATTTCCAACCATATCCACTACC*<br>Sequencing→ GTTAGGTTTTTTGTTATGTTATTG | 60 | 25 | 86 | chr17:56435427-56435508 | 82 | 3 |
| CG25 | PCR→ GGGGAATTTAGGATGGGTATTATAT<br>PCR← *CTCAAAATTACCAACTATTTAAACCATACA*<br>Sequencing→ AATTTAGGATGGGTATTATATT | 60 | 20 | 138 | chr8:141619395-141619528 | 134 | 6 |

**Supplementary table 3 (continuation).**

| Biomarkers | Primers | Singleplex PCR temperature (°C) | Singleplex PCR cycles | Singleplex amplicon length (bp) | Pyrosequenced region (GRCh37/hg19) | Pyrosequenced region length (bp) | Amount of CpG sites analysed |
|---|---|---|---|---|---|---|---|
| CG6 | PCR→ AGTTAGAGTGAGTGGGTAGTA<br>PCR← *CAACCCCCCTTCTACACAAAAACT*<br>Sequencing→ GTGAGTGGGTAGTAAT | 58 | 25 | 86 | chr4:1864164-1864242 | 79 | 3 |
| CG10 | PCR→ TTTAGTGTATTTTTGGGTGTGGTGTTTAT<br>PCR← *ACTATATACAAATAACAAACCAAACTT*<br>Sequencing→ TGTTTTGTATAGTAGAGGTTA | 58 | 25 | 186 | chr16:3534508-3534611 | 104 | 1 |
| CG17 | PCR→ GGAGTTTGGGAAGAAAGTTTTT<br>PCR← *CTACCCATCCTACTACTATCTTCAAAT*<br>Sequencing→ GAAGAAAGTTTTTGTTGTTAG | 58 | 20 | 167 | chr10:130281707-130281864 | 158 | 3 |
| CG7 | PCR→ AGATTAGGGAAGAGTTATTTGGAAAT<br>PCR← *CCTAAAACTAAAAAAAAACCCATTCTACC*<br>Sequencing→ TTTTAATTAGTAAGTTATAGGGAG | 58 | 20 | 125 | chr11:110220584-110220663 | 80 | 1 |
| CG14 | PCR→ ATGGTTTATTATTTTTTTATTTGATAATT<br>PCR← *CACCAATCATTCCTCCAACAAA*<br>Sequencing→ TTGATAATTTAGTATTAGTTTTAG | 58 | 20 | 98 | chr11:110104045-110104122 | 78 | 2 |
| CG22 | PCR→ TTTTTTTATATGGGGATAGGAATGTGATT<br>PCR← *AACCCTTCCCCAACCTCTAATACCA*<br>Sequencing→ GGGGTTTGTTGGTTTT | 58 | 25 | 172 | chr5:1852812-1852926 | 115 | 2 |
| CG23 | PCR→ *GGTAGTGATTATAGTTTGTAGGGGTTTGT*<br>PCR← AACCCTTCCCCAACCTCTAATACC<br>Sequencing← ATCTCCACCAAAAAACACCTAA | 60 | 25 | 135 | chr5:1852792-1852864 | 73 | 1 |
| CG16 | PCR→ *AATAGGGTGGTGAAAGGTAGATAAA*<br>PCR← ACCACAACAACTAAAAACACCTATC<br>Sequencing← ACTAAAAACACCTATCTCC | 58 | 20 | 131 | chr5:140864546-140864667 | 122 | 11 |
| CG19 | PCR→ GTGAGGGGATTGTTGGAAGAG<br>PCR← *TTCCCCCACACCTAACACCCATAT*<br>Sequencing→ AGTAGGAATGTTAATTTGG | 58 | 20 | 183 | chr7:99227410-99227486 | 77 | 4 |
| CG24 | PCR→ *AGAGGAAGTTTTTTTGTTTTATTTTGATA*<br>PCR← ACATCACTATACTTTCACCCTCT<br>Sequencing← CTATACTTTCACCCTCTT | 58 | 20 | 123 | chr4:16723341-16723457 | 117 | 6 |
| CG26 | PCR→ AGGTGGTGGTGTTTGTTTT<br>PCR← *AATACCCACTTAATCCTTAACATCAC*<br>Sequencing→ TTGTTTTAGGTTGGTTTTT | 58 | 25 | 90 | chr22:44319222-44319299 | 78 | 2 |

**Supplementary table 4.** Description of the CpG candidate biomarkers obtained after the epigenome-wide methylation analysis.

| | Methylation EPIC probe ID | Genomic location (GRCh37/hg19) | Gene symbol | Relation to CpG island | CpG island | Regulatory feature |
|---|---|---|---|---|---|---|
| CG1 | cg08942894 | chr15:83563792 | HOMER2 | Opensea | | Body |
| CG2 | cg05445162 | chr2:27730181 | GCKR | Opensea | | Body |
| CG3 | cg11113216 | chr12:132847641 | GALNT9 | Island | chr12:132847640-132847955 | Body |
| CG4 | cg06522913 | chr15:90630711 | IDH2 | Opensea | | Body |
| CG5 | cg14838992 | chr14:100814724 | WARS | Opensea | | Body |
| CG6 | cg06148974 | chr4:1864183 | | Island | chr4:1864182-1864417 | |
| CG7 | cg12737198 | chr11:110220620 | | Opensea | | |
| CG8 | cg03111938 | chr6:169289230 | | Island_Shelf | chr6:169286195-169286688 | |
| CG9 | cg07253636 | chr19:50077766 | NOSIP | Opensea | | 5'UTR |
| CG10 | cg06214087 | chr16:3534568 | NAA60 | Island_Shore | chr16:3534833-3535066 | Body |
| CG11 | cg25942688 | chr2:27016730 | CENPA | Opensea | | 3'UTR |
| CG12 | cg01987330 | chr3:142797310 | | Opensea | | |
| CG13 | cg22880141 | chr1:28160795 | SCARNA1 PPP1R8 | Island_Shelf | chr1:28157221-28157788 | TSS200 |
| CG14 | cg10641001 | chr11:110104096 | RDX | Opensea | | Body |
| CG15 | cg01550272 | chr13:115050881 | UPF3A | Island_Shelf | chr13:115046754-115048034 | Body |
| CG16† | cg03640756 | chr5:140864593 | PCDHG gene cluster | Island | chr5:140864527-140864748 | Body |
| CG17† | cg26024530 | chr10:130281732 | | Opensea | | |
| CG18† | cg22778120 | chr22:45094531 | PRR5 | Island_Shelf | chr22:45097755-45098801 | 5'UTR |
| CG19† | cg15442105 | chr7:99227437 | ZNF498 | Opensea | | Body |
| CG20† | cg04544475 | chr17:56435455 | RNF43 | Opensea | | Body |
| CG21† | cg16639692 | chr5:43037666 | | Island_Shore | chr5:43037259-43037520 | |
| CG22† | cg14969646 | chr1:3148357 | PRDM16 | Island_Shore | chr1:3147845-3148081 | Body |
| CG23† | cg04600077 | chr5:1852839 | | Island_Shore | chr5:1851342-1851564 | |
| CG24‡ | cg14503564 | chr4:16723393 | LDB2 | Opensea | | Body |
| CG25§ | cg18044585 | chr8:141619425 | EIF2C2 | Opensea | | Body |
| CG26‡ | cg23653187 | chr22:44319257 | PNPLA3 | Island_Shore | chr22:44319578-44320513 | TSS1500 |

Regulatory features and relation to CpG island of biomarkers annotated according to the Methylation EPIC Manifest: CpG island: region of at least 200 bp with a CG content > 50% and an observed-to-expected CpG ratio≥0.6; Island-shore: sequences 2 kb flanking the CpG island; Island-shelf: sequences 2 kb flanking shore regions; Opensea: sequences located outside these regions; Body: gene body (intragenic region); TSS200, TSS1500: 200 and 1500 bp upstream the transcription start site, respectively. †Candidate biomarkers derived from the comparison with the external RRBS dataset; ‡candidate biomarkers derived from the NN vs P-AA classification; §candidate biomarker derived from the NN vs D-AA classification.
