## Supplementary information about bioinformatics preprocessing of microarray data, primers and PCR conditions, and final classification rules for "Serum methylation of *GALNT9*, *UPF3A*, *WARS,* and *LDB2* as non-invasive biomarkers for the early detection of colorectal cancer and premalignant adenomas"

### BIOINFORMATICS PREPROCESSING OF METHYLATION MICROARRAY DATA FOR BIOMARKER DISCOVERY

**Quality control.** First, data quality control was assessed based on the internal control probes present on the array. Technical variation, outliers, and potential sources of covariates were explored with multi-dimensional scaling plots and principal component analysis. The distribution of methylation levels was checked across all samples.

**Probe filtering.** Detection p values were computed with the *minfi* package (1), and mean detection p values were examined across all samples to identify any failed sample. Probe filtering was performed with the *wateRmelon* package (2). Probes with a detection p value > 0.01 in at least one sample, probes with a bead count < 3 in at least 1% of samples, and probes that violated any assumption for linear regression model fitting (linearity, homoscedasticity, uncorrelatedness, and normality of the standardized residuals) were discarded. We filtered out probes containing single nucleotide polymorphisms and probes targeting X and Y chromosomes. Cross-reactive probes reported by Pidsley et al. (3) were also removed. No sample was discarded due to quality issues. Probes were annotated to CpG island, genomic regions, and RefSeq genes according to the MethylationEPIC Manifest.[1]

**Normalization and batch effect correction.** The dataset was then normalized using the single-sample out-of-band normalization method (ssNoob) (4, 5) implemented in the *minfi* package. This approach for background and dye bias correction does not rely on biological assumptions, as it is based on the non-specific fluorescence of Infinium I probes. Before differential methylation analysis, data were adjusted for the BeadChip batch effect using the ComBat method (6) implemented in the *sva* package (7).

### PCR CONDITIONS AND PRIMERS FOR BIOMARKER EVALUATION

ABM 2x PCR HotStart Mastermix (Applied Biological Materials, Richmond, Canada) was used for PCR amplification with the following cycling conditions:

- Multiplex PCR: 94°C 10 min, 32 cycles of (94°C 30 s, 58°C 30 s, 72°C 30 s), 72°C 5 min, cool down to 4°C. Ordinary primers were used at a final concentration of 0.6 µM
- Singleplex PCR: 94°C 10 min, 20/25 cycles of (94°C 30 s, 58/60°C 30 s, 72°C 30 s), 72°C 5 min, cool down to 4°C. Primers were used at a final concentration of 0.5 µM. A biotin-labelled primer, either forward or reverse (see Supplementary table 3) was used.

[1] Kasper Daniel Hansen (2016). IlluminaHumanMethylationEPICmanifest: Manifest for Illumina's EPIC methylation arrays. R package version 0.3.0. https://bitbucket.com/kasperdanielhansen/Illumina_EPIC

**FINAL MODELS FOR THE DETECTION OF COLORECTAL ADVANCED NEOPLASIA**

The cut-off value of p for AN classification was determined by the Youden Index method. The final classification models where *p* is the predicted probability of being classified as advanced neoplasia are given by:

*GALNT9/UPF3A* model (AN if p>0.379)

$$logit(p) = -6.223 - 0.0218log(GALNT9 + 1) + 1.270log(UPF3A + 1)$$

*GALNT9*, *UPF3A*, *WARS* and *LDB2* model (AN if p>0.381)

$$logit(p) = -2.332 - 0.006GALNT9 + 0.020UPF3A + 0.002WARS + 0.003LDB2$$

The values of CG3-*GALNT9*, CG15-*UPF3A*, CG5-*WARS* and CG24-*LDB2* correspond to the methylation levels of the CpG sites chr12:132847644, chr13:115050881, chr14:100814747 and chr4:16723393, respectively (GRCh37/hg19 coordinates).
